# Body mass index and childhood symptoms of depression, anxiety, and attention-deficit hyperactivity disorder: a within-family Mendelian randomization study

**DOI:** 10.1101/2021.09.17.21263612

**Authors:** Amanda M Hughes, Eleanor Sanderson, Tim Morris, Ziada Ayorech, Martin Tesli, Helga Ask, Ted Reichborn-Kjennerud, Ole A. Andreassen, Per Magnus, Øyvind Helgeland, Stefan Johansson, Pål Njølstad, George Davey Smith, Alexandra Havdahl, Laura D Howe, Neil M Davies

## Abstract

**Background:** Higher BMI in childhood is associated with emotional and behavioural problems, but these associations may not be causal. Results of previous genetic studies imply causal effects but may reflect influence of demography and the family environment.

**Methods:** This study used data on 40,949 8-year-old children and their parents from the Norwegian Mother, Father and Child Cohort Study (MoBa) and Medical Birth Registry of Norway (MBRN). We investigated the impact of BMI on symptoms of depression, anxiety, and attention-deficit hyperactivity disorder (ADHD) at age 8. We applied within-family Mendelian randomization, which accounts for familial effects by controlling for parental genotype.

**Results:** Within-family Mendelian randomization estimates using genetic variants associated with BMI in adults suggested that a child’s own BMI increased their depressive symptoms (per 5kg/m^2^ increase in BMI, beta=0.26 S.D., CI=-0.01,0.52, p=0.06) and ADHD symptoms (beta= 0.38 S.D., CI=0.09,0.63, p=0.009). These estimates also suggested maternal BMI, or related factors, may independently affect a child’s depressive symptoms (per 5kg/m^2^ increase in maternal BMI, beta=0.11 S.D., CI:0.02,0.09, p=0.01). However, within-family Mendelian randomization using genetic variants associated with retrospectively-reported childhood body size did not support an impact of BMI on these outcomes. There was little evidence from any estimate that the parents’ BMI affected the child’s ADHD symptoms, or that the child’s or parents’ BMI affected the child’s anxiety symptoms.

**Conclusions:** We found inconsistent evidence that a child’s BMI affected their depressive and ADHD symptoms, and little evidence that a child’s BMI affected their anxiety symptoms. There was limited evidence of an influence of parents’ BMI. Genetic studies in samples of unrelated individuals, or using genetic variants associated with adult BMI, may have overestimated the causal effects of a child’s own BMI.

**Funding:** This research was funded by the Health Foundation. It is part of the HARVEST collaboration, supported by the Research Council of Norway. Individual co-author funding: the European Research Council, the South-Eastern Norway Regional Health Authority, the Research Council of Norway, Helse Vest, the Novo Nordisk Foundation, the University of Bergen, the South-Eastern Norway Regional Health Authority, the Trond Mohn Foundation, the Western Norway Regional Health Authority, the Norwegian Diabetes Association, the UK Medical Research Council. The Medical Research Council (MRC) and the University of Bristol support the MRC Integrative Epidemiology Unit.

## Introduction

Children with high body mass index (BMI) have been found to have greater risk of emotional and behavioural problems, including symptoms and diagnoses of depression (Lindberg et al. 2020; Patalay and Hardman 2019; Geoffroy, Li, and Power 2014; Quek et al. 2017) anxiety (Lindberg et al. 2020) and attention-deficit hyperactivity disorder (ADHD) (Cortese and Tessari 2017; Griffiths, Dezateux, and Hill 2011). Prior to the COVID-19 pandemic, prevalence of childhood overweight and childhood obesity, respectively, was 21.3% and 5.7% in Europe(Garrido-Miguel et al. 2019) and 20.1% and 4.3% in Norway(Glavin et al. 2014). The estimated prevalence in Europe of mid-childhood emotional disorders was around 4%(Kovess-Masfety et al. 2016; Sadler et al. 2018) while the global prevalence of child and adolescent ADHD was estimated at 5%(Sayal et al. 2018). These rates may have increased considerably in the wake of the pandemic(Vizard et al. 2020). In this context, there is a clear need to understand the relationship between these factors, but it is not known if child body weight causes emotional or behavioural problems.

High BMI in childhood could affect emotional symptoms through social mechanisms, for example bullying victimization(Puhl et al. 2017). An impact on ADHD has been proposed via sleep disturbance and neurocognitive functioning(Vogel et al. 2015). However, even if children with high BMI are more likely than normal weight children to experience these symptoms, associations may not be causal. Aspects of the family environment may independently affect children’s BMI and their likelihood of developing emotional and behavioural symptoms, for example socioeconomic disadvantage (Russell et al. 2016) and parental mental health(Hope, Micali, et al. 2019; Hope, Pearce, et al. 2019). Some studies have suggested that prenatal maternal obesity may confound associations of childhood BMI with emotional and behavioural symptoms(Sanchez et al. 2018) although the evidence is mixed(Li et al. 2020; Arafat and Minica 2018). Reverse causality is also plausible: depressive, anxiety or ADHD symptoms could cause higher BMI, for instance via disordered eating patterns or decreased physical activity(Blaine 2008; Martins-Silva et al. 2019). To avoid confounding and reverse causation, recent studies have applied Mendelian randomization (MR), a causal inference approach which uses genetic variants as instrumental variables for putative risk factors(Davies, Holmes, and Davey Smith 2018). Results, principally based on adult populations, are consistent with a causal influence of BMI on ADHD(Martins-Silva et al. 2019) and depression(Tyrrell et al. 2019). They are inconclusive for anxiety, reporting both positive(Walter et al. 2015) and negative(Millard et al. 2019) predicted causal effects of body weight.

However, although MR studies avoid classical confounding and reverse causation, they can be vulnerable to other sources of bias. Specifically, estimates from ‘classic’ MR studies – those conducted on samples of unrelated individuals - may be affected by demographic and familial factors(Davies et al. 2019; Morris et al. 2020). Bias can firstly arise from uncontrolled population stratification, where systematic differences in genotype between individuals from different ancestral clusters correlates with differences in environmental or cultural factors. This is an example of gene-environment correlation, which can lead to biased associations of genotypes and phenotypes.

Secondly, indirect genetic effects may exist whereby parental genotype influences a child’s phenotype via environmental pathways, termed ‘dynastic effects’ or ‘genetic nurture’(Kong et al. 2018). Thirdly, assortative mating in the parents’ generation, where parents are more (or less) similar to each other than would be expected by chance, can distort genotype-phenotype associations in the child’s generation. Recent work has suggested that these biases may be especially pronounced for complex social and behavioural phenotypes(Brumpton et al. 2020; L. J. Howe et al. 2022). Previously reported MR estimates of the effect of BMI on emotional and behavioural problems may therefore partly reflect demographic or familial biases rather than a causal influence of BMI. To investigate this, we used a ‘within-family’ Mendelian randomization (within-family MR) design. This approach uses the child’s, mother’s, and father’s genotype data as instruments for the BMI of the child, mother, and father. Within family Mendelian randomization estimates of the effect of the child’s BMI on the outcomes are robust to demographic and family-level biases. We compared within-family MR estimates with estimates from multivariable regression of the child’s outcomes on the child’s, mother’s and father’s reported BMI, and with estimates from ‘classic’ Mendelian randomization (classic MR), in which the child’s genotype data was used to instrument the child’s BMI without controlling for the parents’ genotype.

## Methods

### Study population

The Norwegian Mother, Father and Child Cohort Study (MoBa) is a population-based pregnancy cohort study over 114,500 children, 95,200 mothers, and 75,200 fathers conducted by the Norwegian Institute of Public Health(Magnus et al. 2016). Participants were recruited from all over Norway from 1999-2008, with 41% of all pregnant women invited consenting to participate. The first child was born in October 1999 and the last in July 2009. The cohort now includes over 114,500 children, 95,200 mothers, and 75,200 fathers (for more details see Appendix 1: MoBa study details). As of May 2022, genotype data which had passed quality control filters was available for 76,577 children, 53,358 fathers, and 77,634 mothers. This analysis was restricted to 40,949 mother-father-child ‘trios’ for whom genetic data were available for all three individuals, and at least one questionnaire had been completed.

The numbers of participants excluded are shown in a STROBE flow chart in Appendix 1 – Figure 1. From all records in MoBa (N=114,030, after removing consent withdrawals), participants were excluded if the parents had not completed any of the MoBa questionnaires used in imputation models. Of the 104,915 records remaining, there were 40,949 births for which genetic data were available and had passed QC filters for mother, father, and child (for details see Appendix 1: Genotyping and imputation, and Appendix 1: Genetic quality control). Missing values in phenotypic information for these participants were estimated using multiple imputation (details in Appendix 1: Multiple imputation). Related participants were retained, but all models were clustered by genetic family ID derived using KING software(Manichaikul et al. 2010). This genetic family ID groups first, second, and third-degree relatives (i.e., siblings in the parental generation and their children as well as nuclear families), in this way accounting for non-independence of observations.

### Measures

Children’s BMI was calculated from height and weight values reported by mothers when the children were 8 years old. Maternal pre-pregnancy BMI was calculated from height and weight reported at ∼17 weeks gestation. Father’s BMI was calculated from self-reported height and weight at ∼17 weeks gestation. This information was missing from around 60% of fathers, and in these cases the mother’s report of the father’s height and weight was used instead (observed values of BMI from the two sources were correlated at 0.98). Values of height and weight more than 4 standard-deviations from the mean were treated as outliers and coded to missing.

Depressive, anxiety, and ADHD symptoms were reported by the mother when the child was 8 years old using validated measures. For depressive symptoms, the 13-item Short Mood and Feelings Questionnaire (SMFQ) was used, for anxiety symptoms the 5-item Short Screen for Child Anxiety Related Disorders (SCARED)(Birmaher et al. 1999) and for ADHD symptoms the Parent/Teacher Rating Scale for Disruptive Behaviour Disorders (RS-DBD) (total score and subdomain scores for inattention and hyperactivity)(Silva et al. 2005). Prorated summary scores were calculated for individuals with at least 80% of item-level information. Full details of all questions asked in MoBa are available at https://mobawiki.fhi.no/mobawiki/index.php/Questionnaires.

Blood samples were obtained from both parents during pregnancy and from mothers and children (umbilical cord) at birth. Details of genotyping and genetic quality control are described in Appendix 1: Genotyping and imputation and Appendix 1: Genetic quality control. Polygenic scores (PGS) for BMI were calculated using SNPs previously associated in GWAS with BMI at p<5.0×10^−8^ and weighted using the individual SNP-coefficients from the GWAS. We first constructed a PGS based on the largest existing GWAS of BMI in adults(Yengo et al. 2018) Since genetic influences on BMI in childhood and adulthood differ(Silventoinen et al. 2016) we also constructed a PGS based on a GWAS of body size in childhood as recalled by adult participants of UK Biobank(Richardson et al. 2020). These SNPs have been shown in external validation samples to predict BMI in childhood better than SNPs associated with adult BMI(Richardson et al. 2020; Brandkvist et al. 2020). From the full GWAS results, we excluded SNPs not available in MoBa, then used the TwoSampleMR package (Hemani et al. 2018) to identify SNPs independently associated with BMI (with a clumping threshold of r=0.01, LD=10,000kb) at p<5.0×10^−8^. This left 954 SNPs associated with adult BMI, and 321 associated with childhood body size. Full details of SNPs included in both PGSs are provided in Supplementary Files 1a and 1b. Equivalent PGSs were derived for depression and ADHD based on SNPs previously associated with these conditions at p<5.0×10^−8^ in GWAS(Wray et al. 2018; Demontis et al. 2019). This was not possible for anxiety, due to few known SNPs associated with these traits at p<0.05×10^−8^. Details of the SNPs in the depression and ADHD PGSs are provided in Supplementary Files 1c and 1d.

### Statistical analysis

Among trios with genetic data, multiple imputation by chained equations was performed in STATAv16 to estimate missing phenotypic information (details in Appendix 1: Multiple imputation of phenotypes). We used non-genetic linear regression, classic MR, and within-family MR to estimate the effects of the child’s BMI on the following outcomes: depressive, anxiety, and ADHD symptoms, and subdimensions of ADHD (inattention and hyperactivity). Non-genetic regression models were adjusted for child’s sex, year of birth, mother’s and father’s BMI, and likely confounders of observational associations: mother’s and father’s educational qualifications, mother’s and father’s depressive/anxiety symptoms (using selected items from the 25-item Hopkins Checklist(Hesbacher et al. 1980)) and ADHD symptoms (from the 6-item adult ADHD self-report scale(Kessler et al. 2005)), mother’s and father’s smoking status during pregnancy, and maternal parity at the child’s birth. For comparability, these models also included all covariates included in genetic models: genotyping centre, genotyping chip, and 20 principal components of ancestry for the child, mother, and father (for detailed information on principal components see Appendix 1: Genetic quality control). All MR models were conducted with two-stage least squares instrumental-variable regression using Stata’s ivregress, with F-statistics and R^2^ values obtained using ivreg2. Classic MR models, which do not account for parental genotype, used the child’s own PGS but not those of the parents to instrument the child’s BMI. Within-family MR models were multivariable MR models, in which we used PGSs for all members of a child-mother-child trio to instrument the BMI of all three individuals (model equations are provided in Appendix 1: Model equations). Classic and within-family MR models were adjusted for the child’s sex and year of birth, and the genotyping centre, genotyping chip, and the first 20 principal components of ancestry for the child, mother, and father. Given skew in outcomes variables, all models used robust standard errors (Stata’s vce option) and thus made no assumptions about the distribution of outcomes. We report two sets of results, in which either the adult BMI GWAS, or the childhood body size GWAS, was used to create the BMI PGS for the child, mother, and father. Z tests of difference were used to formally compare the classic MR and within-family MR estimates. To assess the extent of assortative mating in the parental generation based on phenotype data, we ran linear regression models of standardized paternal BMI, depressive symptoms, and ADHD symptoms on standardized maternal BMI, depressive symptoms, and ADHD symptoms. We then regressed paternal polygenic scores for BMI, depression, and ADHD on maternal polygenic scores for BMI, depression, and ADHD. All models investigating assortative mating adjusted for both parents’ principal ancestry components and genotyping covariates. We did not examine correlations with polygenic scores for anxiety, due to few known SNPs associated with these traits at p<0.05×10^−8^. All statistical tests were two-tailed.

### Sensitivity analyses

To check sensitivity of results to outliers, all analyses were repeated using log-transformed versions of outcome measures (as all symptoms scales began at 0, we added 1 to scores before log-transforming). Genetic studies designed to assess causation can be biased by horizontal pleiotropy(Davies, Holmes, and Davey Smith 2018). This is when genetic variants in a polygenic score influence the outcome via pathways which do not involve the exposure. Pleiotropic effects can inflate estimated associations, or bias estimates towards the null. Methods have been developed to test for the presence of horizontal pleiotropy by comparing SNP-specific associations of exposures and outcomes, although these tests themselves rest on assumptions(Hemani, Bowden, and Davey Smith 2018). We therefore performed additional robustness checks based on associations of individual SNPs included in the polygenic scores with BMI in the GWAS, and associations of the same SNPs with each outcome in MoBa. It was not computationally feasible to include individual SNPs in the imputation models, so SNP-outcome associations in MoBa were calculated using unimputed SNP data with imputed outcome data. For robustness checks of classic MR models, SNP-outcome associations were adjusted for the child’s sex and birth year, and the genotyping centre, genotyping chip, and ancestry principal components of the child, mother, and father. For robustness checks of within-family MR models, SNP-outcome associations were adjusted for the child’s sex and birth year, mother’s and father’s genotype, and the genotyping centre, genotyping chip, and principal components of the child, mother, and father. We conducted inverse-variance weighted, MR-Median, MR-Mode, and MR-Egger regression in STATAv16 with the MRRobust package(Spiller, Davies, and Palmer 2019). A non-zero intercept from an MR-Egger model indicates presence of horizontal pleiotropy. We repeated main analyses without using imputed data in the sample of participants who had full genetic, exposure, outcome, and covariate data. To explore nonlinearities in associations of BMI with depression, anxiety, and ADHD symptoms, we ran non-genetic models with the child’s BMI divided into quintiles. Finally, MR models were run with additional adjustment for parental education. Attenuation of classic MR estimates in these models would be consistent with confounding by aspects of the family environment linked to parental education.

## Results

This analysis was restricted to 40,949 mother-father-child ‘trios’ for whom genetic data were available for all three individuals, and at least one questionnaire had been completed. To assess whether participants included in the analytic sample (N=40,949) differed from the rest of the MoBa sample (N=72,742), we conducted t-tests and chi-squared tests for key characteristics at birth, BMI, and outcomes using unimputed data. There were modest differences, described in Appendix 1: Comparison of analytic sample and excluded participants. BMI did not differ for mothers, fathers or children, but children in the analytic sample had slightly lower depressive symptoms (mean SMFQ=1.81 vs 1.91), anxiety symptoms (mean SCARED=1.04 vs 1.00) and ADHD symptoms (mean RS-DBD ADHD=8.4 vs 8.7). Descriptive characteristics of the full MoBa sample are in Appendix 1 - Table 1.

Descriptive statistics of the analytic sample after multiple imputation is presented in Table 1. The mean BMI for children was 16.3 (SD=2.0), for mothers 24.0 (SD=4.1), and for fathers 25.9 (SD=3.2). Corresponding descriptive characteristics from unimputed data are included in Appendix 1 - Table 2. Both polygenic scores used to instrument BMI were strong instruments, even when used in within-family models. For the adult BMI PGS, conditional first-stage F-statistics for children, mothers, and fathers were 718.7, 1338.2, and 1272.5. The conditional R^2^ showed that the score explained 1.7%, 3.2%, and 3.0% of the variation in BMI for children, mothers, and fathers respectively. For the childhood body size PGS, conditional first-stage F-statistics were 919.8, 1071.8 and 960.2 for children, mothers, and fathers, with the scores explaining 2.2%, 2.6% and 2.3% of the variation in BMI. The correlation of the polygenic scores for adult BMI and for childhood body size was 0.38 for children, 0.36 for mothers and 0.37 for fathers.

**Table 1.** Descriptive Characteristics of Analytic Sample (N=40,949)^a^.

| **Continuous variables** |  | **mean** | **SD** |
| --- | --- | --- | --- |
| Maternal age at child's birth (years) |  | 30.2 | 4.4 |
| Paternal age at birth (years) |  | 32.6 | 5.1 |
| Maternal depressive/anxiety symptoms, Hopkins Symptoms Checklist-25 (SCL-25)<sup>b</sup> |  | 1.2 | 1.9 |
| Paternal depressive/anxiety symptoms, Hopkins Symptoms Checklist-25 (SCL-25)<sup>c</sup> |  | 1.1 | 2.1 |
| Maternal ADHD symptoms: adult ADHD self-report scale<sup>d</sup> |  | 6.7 | 3.4 |
| Paternal ADHD symptoms: adult ADHD self-report scale<sup>e</sup> |  | 8.3 | 3.1 |
| Maternal pre-pregnancy BMI (kg/m<sup>2</sup>) |  | 24.0 | 4.1 |
| Paternal BMI (kg/m<sup>2</sup>) |  | 25.9 | 3.2 |
| Child's BMI at age 8 (kg/m<sup>2</sup>) |  | 16.3 | 2.0 |
| Child depressive symptoms age 8: Short Mood and Feelings Questionnaire (SMFQ)<sup>f</sup> |  | 1.9 | 2.5 |
| Child anxiety symptoms age 8: Screen for Child Anxiety Related Disorders (SCARED)<sup>g</sup> |  | 1.0 | 1.2 |
| Child ADHD symptoms age 8: Parent/Teacher Rating Scale for Disruptive Behaviour Disorders (RS-DBD)<sup>h</sup> |  | 8.6 | 7.2 |
| Child ADHD symptoms (inattention) age 8: Parent/Teacher Rating Scale for Disruptive Behaviour Disorders (RS-DBD)<sup>i</sup> |  | 5.0 | 4.1 |
| Child ADHD symptoms (hyperactivity) age 8: Parent/Teacher Rating Scale for Disruptive Behaviour Disorders (RS-DBD)<sup>j</sup> |  | 3.6 | 3.9 |
| **Categorical variables** | **Category** | **%** |  |
| Child's sex | Male | 51.1 |  |
|  | Female | 48.9 |  |
| Maternal educational qualifications | 9-year elementary education | 2.0 |  |
|  | Up to 2 years further education | 4.1 |  |
|  | Further education: vocational | 12.2 |  |
|  | Further education: general studies, sixth form | 14.9 |  |
|  | Higher education: college/university, up to 4 years | 42.8 |  |
|  | Higher education: college/university, over 4 years | 24.0 |  |
| Paternal educational qualifications | 9-year elementary education | 3.4 |  |
|  | Up to 2 years further education | 5.6 |  |
|  | Further education: vocational | 25.1 |  |
|  | Further education: general studies, sixth form | 12.9 |  |
|  | Higher education: college/university, up to 4 years | 28.4 |  |
|  | Higher education: college/university, over 4 years | 24.4 |  |
| Maternal parity at child's birth | 0 | 46.8 |  |
|  | 1 | 35.7 |  |
|  | 2 | 14.0 |  |
|  | 3+ | 2.7 |  |
|  | 4+ | 0.7 |  |
| Mother's marital status at birth | Married/registered partner | 97.4 |  |
|  | single | 2.6 |  |
| Mother's smoking status during pregnancy | never | 51.0 |  |
|  | Stopped before week 17 | 42.0 |  |
|  | Currently, sometimes | 2.4 |  |
|  | Currently, daily | 4.5 |  |

<sup>a</sup>The reasons for exclusions and numbers in each case are shown in Appendix 1 - Figure 1. Missing data in BMI, outcomes and covariates was imputed using multiple imputation by chained equations. Descriptive
statistics for the unimputed data are shown in Appendix 1. <sup>b</sup>Based on 5 items. Possible range: 0-15. <sup>c</sup>Based on 8 items. Possible range: 0-24. <sup>d</sup>Possible range: 0-24. <sup>e</sup>Possible range: 0-24. <sup>f</sup>Possible range: 0-10. <sup>g</sup>Possible range: 0-54. <sup>h</sup>Possible range: 0-27. <sup>i</sup>Possible range: 0-27.

### Associations of BMI with depressive, anxiety, and ADHD symptoms at age 8

#### Depressive symptoms (SMFQ)

In adjusted non-genetic regression models (Figure 2, Appendix 1 - Table 3), children’s higher BMI at age 8 was associated with slightly higher depressive symptoms. Per 5kg/m^2^ increase in BMI, SMFQ score was 0.05 standard deviations (SD) higher (95% CI: 0.01,0.09, p=0.02). Classic MR using the adult BMI PGS suggested that for each 5kg/m^2^ increase in the child’s BMI, the child’s SMFQ score increased by 0.45 SD (95% CI: 0.26,0.64, p<0.001). Within-family MR using the adult BMI PGS also provided some evidence for an effect (beta: 0.26 SD, 95% CI: -0.01,0.52, p=0.06), but the within-family MR estimate was less precise (70% as precise as the classic MR estimate, and 15% as precise as the OLS estimates, from the ratio of standard errors). A z test for the difference (p=0.26) indicated that the within-family MR estimate was consistent with the classic MR estimate. Using the childhood body size PGS (Figure 3, Appendix 1 - Table 4) there was little evidence that a child’s own BMI affected their depressive symptoms from either classic MR (beta: 0.08 (95% CI: -0.07,0.22, p=0.29) or within-family MR (beta: 0.02 (95%CI: -0.20,0.23, p=0.88). In summary, evidence for an effect of childhood BMI on depressive symptoms was strongest using the genetic variants for adult BMI.

**Figure 1.**
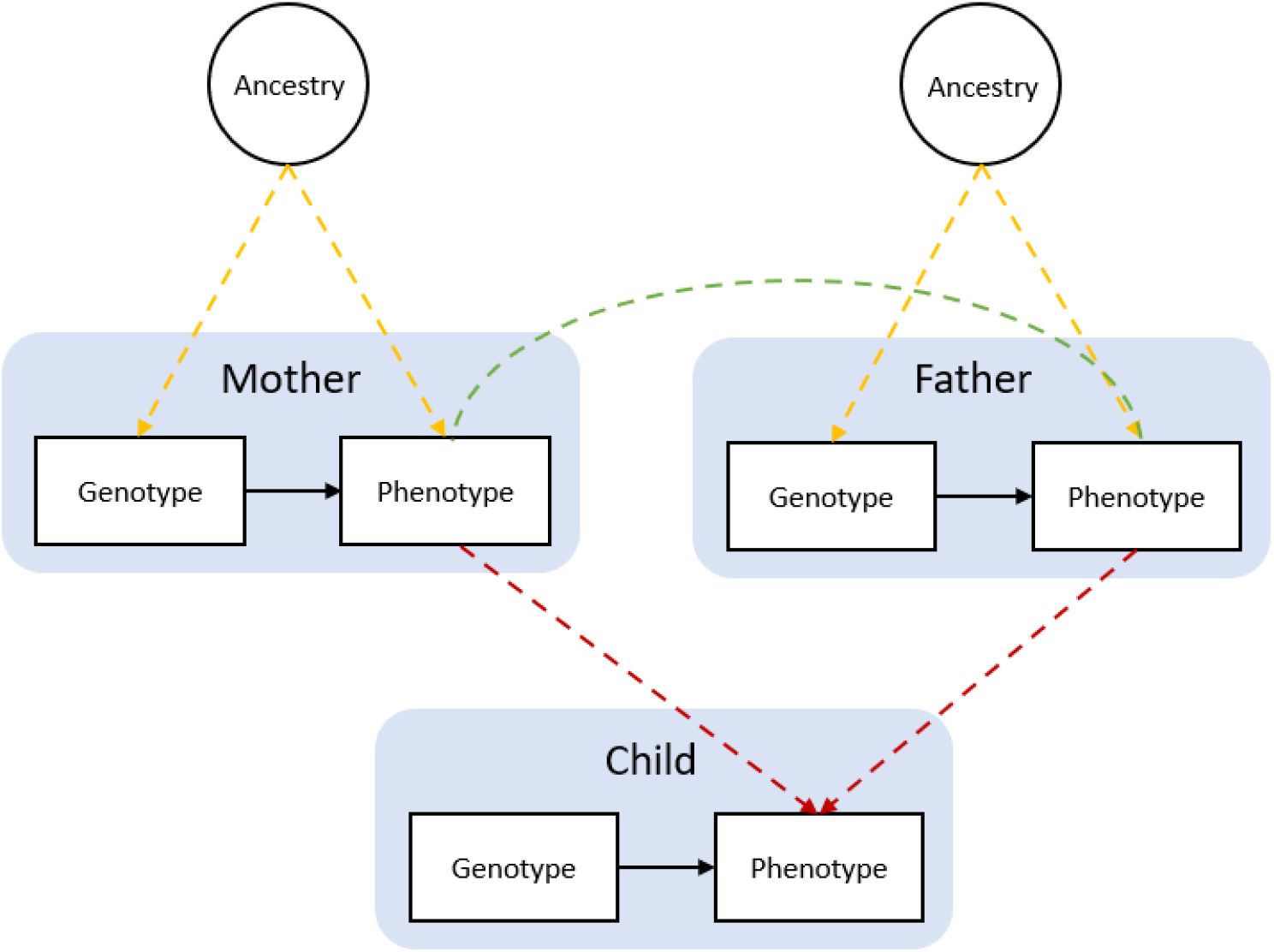
Bias in Mendelian randomization studies which do not account for parental genotype. Figure 1 is reproduced from Figure 1 Morris et al., 2020. Population stratification due to ancestral differences (yellow lines), dynastic effects (red lines), and assortative mating (green line). In within-family Mendelian randomization, parental genotype is controlled for, so effect estimates for the influence of child’s genotype on child phenotypes are unbiased by these processes.

**Figure 2.**
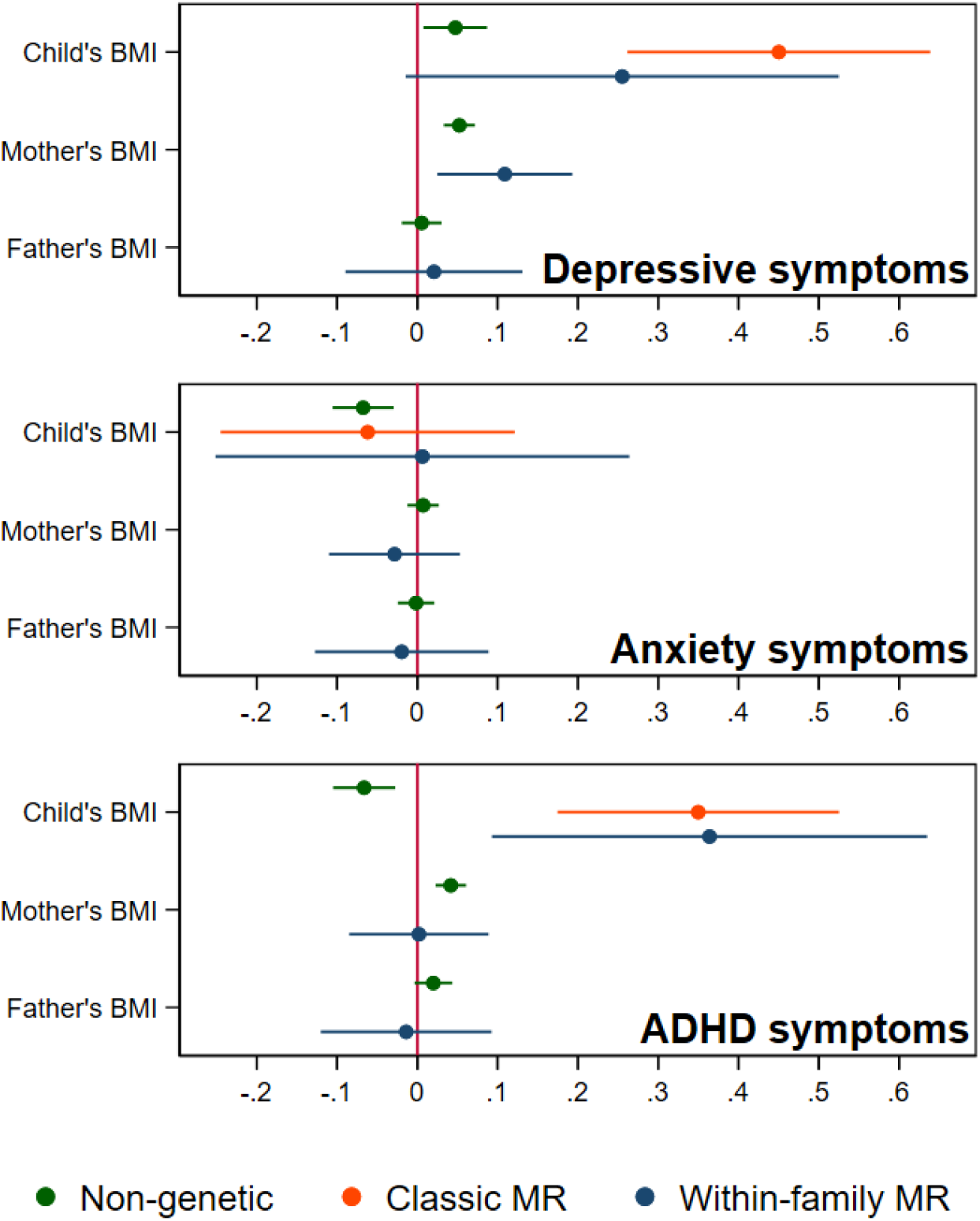
BMI and child’s depressive, anxiety, and ADHD symptoms, using a polygenic score for adult BMI. Coefficients represent standard-deviation change in outcomes per 5kg/m^2^ increase in BMI.

**Figure 3.**
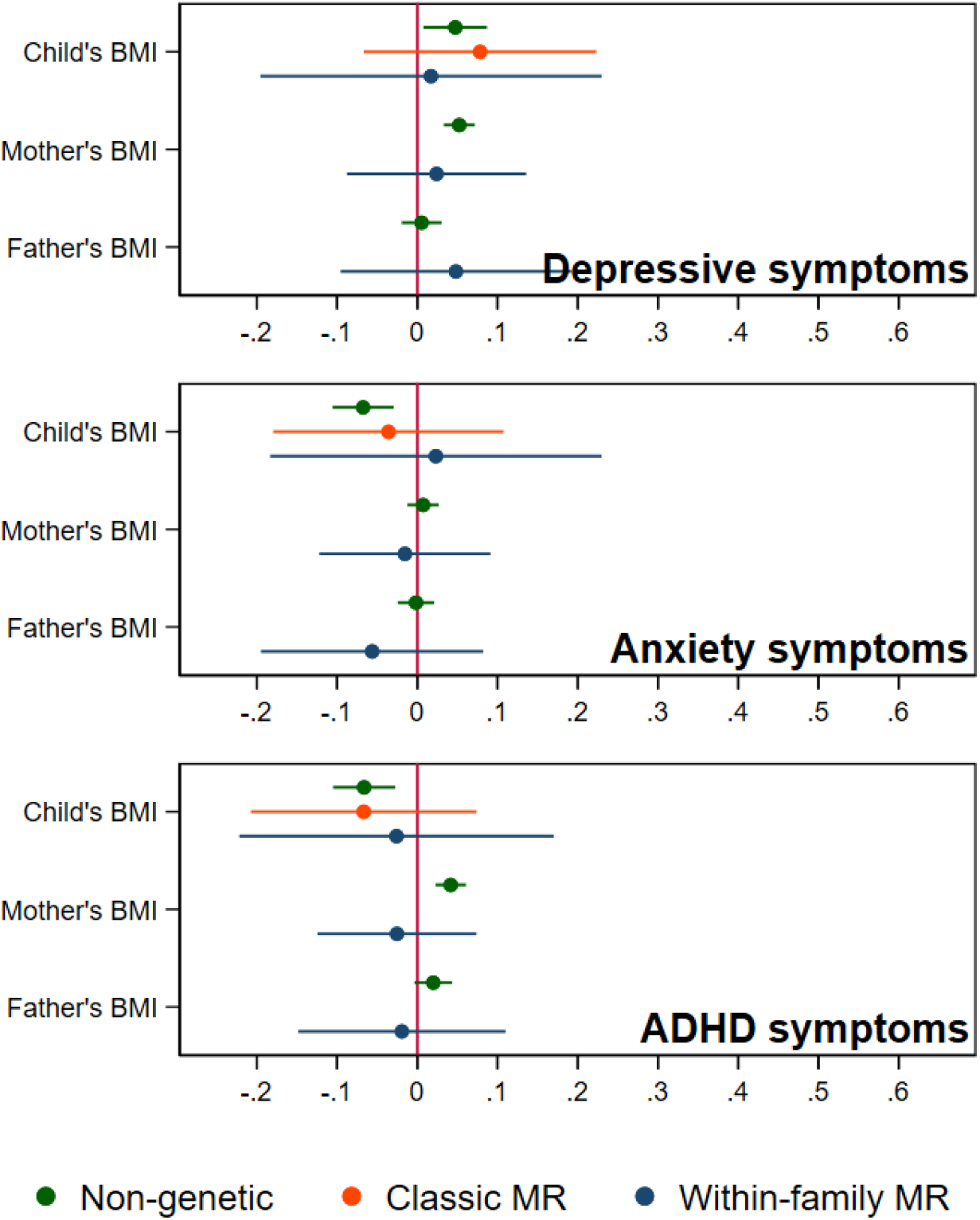
BMI and child’s depressive, anxiety, and ADHD symptoms, using a polygenic score for childhood body size. Coefficients represent standard-deviation change in outcomes per 5kg/m^2^ increase in BMI.

#### Anxiety symptoms *(SCARED)*

In non-genetic models (Figure 2, Appendix 1 - Table 3), each 5kg/m^2^ increase in BMI was associated with a 0.07 SD lower (95% CI: -0.11, -0.03, p=0.001) SCARED score. Using the adult BMI PGS, there was little evidence for an effect from classic MR (beta: -0.06, 95% CI: -0.25,0.12, p=0.51), or within-family MR models (beta: 0.01, 95% CI: -0.25,0.29, p=0.96). Again, the within-family MR estimate was less precise than the classic MR estimate (68% as precise), or the OLS estimate (15% as precise), and the classic and within-family MR estimates were consistent (p=0.54). Using the childhood body size PGS (Figure 3, Appendix 1 - Table 4), MR estimates were similar (classic MR beta: -0.04, 95%: - 0.18,0.11, p=0.62, within-family MR beta: 0.02, 95%CI: -0.18,0.22, p=0.83). In summary, there was little evidence from any genetic model that childhood BMI affects anxiety symptoms.

#### ADHD symptoms (RS-DBD)

In non-genetic models (Figure 2, Appendix 1 - Table 3) children’s BMI was negatively associated with ADHD symptoms after adjusting for confounders. Per 5kg/m^2^ increase in BMI, ADHD symptoms from the RS-DBD were 0.07 SD lower (95% CI: -0.11,-0.03, p=0.001), with similar associations observed for the inattention or hyperactivity subscales (Figure 2, Appendix 1 – Table 3). Using the adult BMI PGS there was evidence from both classic and within-family MR models of a positive association of BMI and ADHD. In the classic MR model ADHD symptoms were 0.35 SD higher (95% CI: 0.17,0.53, p<0.001) per 5kg/m^2^ increase in BMI; the within-family MR estimate, at 0.36 SD (CI: 0.09,0.63, p=0.009) was almost identical (p for difference=0.95). A similar pattern was seen with the inattention and hyperactivity subscales (Figure 2, Appendix 1 - Table 3). The within-family MR estimate was again the least precise (65% as precise as the classic MR estimate, 14% as precise as the non-genetic estimate). Using the childhood body size PGS (Figure 3, Appendix 1 - Table 4) there was little evidence of an association from either classic MR (beta: -0.07, 95%CI: -0.21,0.07, p=0.35) or within-family MR models (beta: -0.03, 95%CI: -0.22,0.17, p=0.80). Thus, as for depressive symptoms, evidence for an effect of childhood BMI on ADHD symptoms was inconsistent and only detected using the adult BMI polygenic score.

### Association of mother’s and father’s BMI with child’s symptoms

In non-genetic models which adjusted for the child’s BMI as well as covariates, the mother’s BMI was associated with slightly more depressive symptoms in the child (the child’s SMFQ score was 0.05 SD higher (95% CI: 0.03,0.07, p<0.001), per 5kg/m^2^ increase in maternal BMI). Maternal BMI was also associated with more ADHD symptoms in the child: the child’s RS-DBD score was 0.04 S.D. higher (95%CI: 0.02,0.06, p<0.001) per 5kg/m^2^ increase in maternal BMI, with similar associations for inattention and hyperactivity subscales. No such associations were seen with paternal BMI.

Within-family MR models also provide estimates for the effect of factors linked to maternal and paternal BMI on child outcomes, conditional on the child’s own BMI. However, compared to within-family MR estimates for the child’s own genotype, the interpretation of parental estimates differs. Like classic MR estimates for the child’s BMI, within-family MR estimates for each parent’s BMI will capture the causal effect of the parent’s BMI on the child’s outcome, but can also reflect residual population stratification and assortative mating in the parents’ generation or earlier. For an unbiased estimate of parental effects, we would need to account for grandparental genotype. Within-family MR models provided inconsistent consistent evidence that maternal BMI affected the child’s depressive symptoms: using the adult BMI PGS (Figure 2, Appendix 1 - Table 3), estimates suggested that higher maternal BMI increased depressive symptoms in the child (0.11 SD higher SMFQ score (95% CI: 0.02,0.19, p=0.01) per 5kg/m^2^ increase in maternal BMI), but within-family MR models using the childhood body size PGS did not (Figure 3, Appendix 1 - Table 4). There was little evidence from within-family MR of other maternal or paternal effects on the child’s emotional or behavioural outcomes.

In the parents’ generation, phenotypes were associated within parental pairs, consistent with assortative mating on these traits (Appendix 1 - Table 5). Adjusted for ancestry and other genetic covariates, maternal and paternal BMI were positively associated (beta: 0.23, 95%CI: 0.22,0.25, p<0.001), as were maternal and paternal depressive symptoms (beta: 0.18, 95%CI: 0.16,0.20, p<0.001), and maternal and paternal ADHD symptoms (beta: 0.11, 95%CI: 0.09,0.13, p<0.001). Consistent with cross-trait assortative mating, there was an association of mother’s BMI with father’s ADHD symptoms (beta: 0.03, 95%CI: 0.02,0.05, p<0.001) and mother’s ADHD symptoms with father’s depressive symptoms (beta: 0.05,95%CI: 0.05,0.06, p<0.001). Phenotypic associations can reflect the influence of one partner on another as well as selection into partnerships, but regression models of paternal polygenic scores on maternal polygenic scores also pointed to a degree of assortative mating. Adjusted for ancestry and genotyping covariates, there were small associations between parents’ BMI polygenic scores (beta: 0.01, 95%CI: 0.00,0.02, p=0.02 for the adult BMI PGS, and beta: 0.01, 95%CI: 0.00,0.02, p=0.008 for the childhood body size PGS), and of the mother’s childhood body size PGS with the father’s ADHD PGS (beta: 0.01, 95%CI: 0.00,0.02, p=0.03). We did not detect associations with pairs of other polygenic scores, which may be due to insufficient statistical power.

### Sensitivity analyses

Analyses using log-transformed versions of the outcomes (Appendix 1 - Tables 6 and 7) were consistent with main results. Robustness checks based on comparing associations of individual SNPs with BMI in the GWAS and with children’s outcomes in MoBa (Appendix 1 - Tables 6 and 7) were consistent with the main results. MR-Egger models found little evidence of horizontal pleiotropy, although MR-Egger estimates were imprecise (Appendix 1 - Tables 8 and 9). Results of analyses using the complete-case sample were qualitatively similar to results using imputed data (Appendix 1 - Tables 10 and 11). In non-genetic models where the child’s BMI was divided into quintiles (Appendix 1 – Table 12), there was little evidence of nonlinear associations. With additional adjustment for parental education, point estimates for depressive and ADHD symptoms in classic MR models were closer to the null, but confidence intervals substantially overlapped (Appendix 1 – Tables 13 and 14).

## Discussion

In a large cohort of Norwegian 8-year-olds, higher childhood BMI was phenotypically associated with slightly more depressive symptoms, but fewer anxiety symptoms and ADHD symptoms. Genetic analyses using the adult BMI PGS suggested that higher BMI in childhood increased symptoms of both depression and ADHD. This was clearest in classic MR models, but also suggested by within-family MR models, whose precision is lower but which account for parental genotype. Compared to associations from non-genetic models, effect sizes for depression and ADHD from genetic models based on the adult BMI PGS were larger. However, these estimates were less precise, and confidence intervals for the classic MR and within-family MR estimates substantially overlapped for all outcomes. The childhood body size PGS explained more variation in children’s BMI than the adult BMI PGS did, consistent with other studies(Richardson et al. 2020; Brandkvist et al. 2020), while the adult BMI PGS explained more variation in maternal and paternal BMI. Genetic analyses which used the childhood body size SNPs provided little evidence that the child’s BMI affected their depressive or ADHD symptoms outcomes. This suggests that genetic variation associated with adult BMI has a greater impact on these outcomes than genetic variation associated with recalled childhood body size. This is consistent with the moderate correlation observed between the two polygenic scores, indicating that they capture both overlapping and unique variation. Our results may therefore reflect differences in how each set of SNPs relate to traits other than childhood BMI which are relevant to a child’s depressive and ADHD symptoms. Nevertheless, within-family MR estimates using the childhood body size PGS were still consistent with small effects of the child’s BMI on all outcomes, with upper confidence limits around a 0.2 standard-deviation increase in each outcome per 5kg/m^2^ increase in BMI. There was little evidence that maternal or paternal BMI affected a child’s ADHD or anxiety symptoms. In within-family MR models using the adult BMI PGS, but not the childhood body size PGS, maternal BMI was positively associated with children’s depressive symptoms. This is consistent with a causal impact of the mother’s recent BMI but not their BMI in childhood, but it may also reflect family-level biases from previous generations.

The positive association between BMI and depressive symptoms in non-genetic models accords with previous observational studies(Lindberg et al. 2020; Patalay and Hardman 2019; Quek et al. 2017; Geoffroy, Li, and Power 2014). The inverse association between BMI and anxiety symptoms in non-genetic models contrasts with the results of a recent study, in which Swedish 6-17 year olds receiving treatment for obesity had a greater likelihood of a diagnosis or prescription for anxiety disorder compared to controls(Lindberg et al. 2020). The discrepancy may reflect confounding (we adjusted for more factors, including parental BMI), age of the participants (children in our study were younger) or differences in the outcome or exposure, since we considered anxiety symptoms rather than diagnosis, and a continuous BMI measure rather than obesity. However, anxiety symptoms in our sample were not raised in the top BMI quintile. Another difference concerns the population: children receiving obesity treatment may be more likely than other children with obesity to experience anxiety symptoms or to receive a diagnosis. The inverse association between BMI and ADHD symptoms in non-genetic models contrasts with previous reports of positive or null associations with obesity, which typically adjusted for fewer confounders(Cortese and Tessari 2017; Nigg et al. 2016). Since previous studies have found more evidence of an association in adults than children, and often considered ADHD diagnoses rather than symptoms, the discrepancy may also point to age-varying associations, or to different influences on likelihood of diagnosis compared to parent-reported symptoms (Nigg et al. 2016; Cortese and Tessari 2017).

For depressive symptoms and ADHD, classic and within-family MR estimates using the adult BMI PGS were larger than estimates from non-genetic models. Horizontal pleiotropy, which we could not rule out, could have inflated MR estimates. It could also help explain the discrepancy in results using the adult BMI and childhood body size polygenic scores, if SNPs in the adult BMI polygenic score have a greater impact on depressive or ADHD symptoms via pathways independent of childhood BMI. We found little evidence of pleiotropy using MR-Egger estimators, but the power to detect pleiotropy with this method is low. Additionally, classic MR estimates may be inflated by demographic and familial factors, but within-family MR estimates for effects of a child’s own BMI are robust to these factors. For depressive symptoms, the within-family MR estimate was closer to the non-genetic estimate than the classic MR estimate, which may reflect bias in the classic MR estimate due to demographic and familial factors. At the same time, the within-family MR estimate was imprecise, and confidence limits consistent with a substantial effect of children’s BMI on depressive symptoms. For ADHD, point estimates from the classic MR and within-family MR models using the adult BMI PGS were very similar, and both statistically distinguishable from the null. These results therefore accord with a recent study which accounted for family-level biases by using dizygotic twin pairs, obtaining between-family and within-family estimates for the effect of BMI on ADHD symptoms (Liu et al., 2020). Using a PGS of SNPs associated with adult BMI, within-family analysis found a 0.07 S.D. increase in ADHD symptoms at age 8 per S.D. increase in BMI PGS, which was consistent with the between-family estimate. The between-family estimate was attenuated by adjustment for parental education, suggesting an influence of family-level processes. In our study, classic MR estimates for depressive and ADHD symptoms which adjusted for parental education were consistent with the main results, with largely overlapping confidence intervals, although point estimates were closer to the null. Thus, our results are also consistent with an influence of demographic or family-level effects, and with earlier evidence that such processes impact the relationship between BMI and ADHD (Chen et al. 2014; Geuijen et al. 2019).

Several sources of genetic familial bias may have influenced classic MR estimates of the impact of the child’s own BMI. Firstly, frequencies of BMI-associated variants may differ between sub-populations in a similar manner to environmental influences on emotional or behavioural functioning (population stratification). Such gene-environment correlation can inflate estimates from classic MR models, but are unlikely to affect within-family MR models, where ancestry is fully controlled for via parental genotypes. Although we included principal components of ancestry in all models, residual population stratification may nevertheless have influenced the classic MR results. Secondly, there may be indirect effects of parental BMI via the family environment (dynastic effects, or genetic nurture). This could explain the association of maternal BMI with children’s depressive symptoms in the within-family MR model using the adult BMI PGS. In observational studies, maternal pre-pregnancy obesity is linked with children’s risk of emotional disorders and ADHD(Sanchez et al. 2018). Although mechanisms are not well understood, an *in utero* effect on children’s neurodevelopment of metabolic correlates of obesity has been proposed(Edlow 2017). Our within-family MR results suggest that previously reported associations of maternal BMI with a child’s ADHD are not causal, but are consistent with an effect on the child’s depressive symptoms. This could reflect an impact of maternal BMI later in the child’s life. A well-documented ‘wage penalty’ exists for high BMI(L. D. Howe et al. 2019), especially for women(Bozoyan and Wolbring 2018) reflecting social consequences of obesity being a stigmatized condition(Giel et al. 2010). High BMI in adulthood is also linked to worse mental health, with stronger associations for women again pointing to gendered social processes(Rubino et al. 2020). Maternal BMI may therefore influence children’s emotional and behavioural problems via economic consequences, or via maternal mental health, throughout childhood. However, while our results are consistent with an influence of maternal BMI on child’s depressive symptoms, these results should be interpreted with caution. In contrast to estimated effects for the child’s BMI, where controlling for parental genotype is likely to eliminate familial biases, estimated maternal and paternal effects from within-family MR models may have been impacted by familial biases in previous generations. Adjustment for grandparental genotype would be required to obtain similarly unbiased estimates for the parents. Thirdly, people with high BMI may be more likely to partner with people with emotional or behavioural conditions (cross-trait assortative mating). Over generations, this would induce an association of not only the phenotypes but of associated genetic variants. We found some genomic evidence of assortative mating for BMI, and cross-trait assortative mating between BMI and ADHD, but not between other traits. However, associations between polygenic scores, which only capture some of the genetic variation associated with these phenotypes, may not capture the full extent of genetic assortment on these traits.

Despite a high participation rate, MoBa is not perfectly representative, and selection biases linked to participation could have affected our results. The current analyses were restricted to families with complete genetic data and at least some relevant questionnaire data. These families were found to have slightly more years of education than the wider MoBa sample, and the children to score slightly lower for depressive, anxiety, and ADHD symptoms. Reflecting the requirement of genetic data for fathers, single mothers were under-represented. Analyses were restricted to individuals of European ancestry, with polygenic scores based on results of GWAS which were also restricted to individuals of European ancestry. Consequently, our results may not be generalisable to other populations. Outcomes were based on mother-reported symptoms of depression, anxiety disorders and ADHD, and estimates based on diagnoses may have differed. However, a child’s sociodemographic characteristics can influence their likelihood of diagnosis independently of symptoms(Thompson, Wilkinson, and Woo 2020), indicating that such an approach is not always preferable. BMI measurements were based on reported height and weight, so reporting bias may have influenced relationships. In many families, fathers’ BMI was based on height and weight reported by the mothers. However, these measures were very highly correlated with father’s self-reports, so additional measurement error is unlikely to have greatly affected our results for father’s BMI. Due to attrition, a substantial proportion of values for the child’s BMI and outcomes were imputed, and we cannot be sure that observations were missing at random conditional on variables included in imputation models. Effects of parental BMI may be time-varying, for example a parent’s own BMI during childhood could influence their child independent of the parent’s later BMI. We could not explore these effects because information on parent’s childhood BMI was not available. Within-family MR may still be affected by horizontal pleiotropy, and recent genetic work points to genetic overlap between BMI and psychiatric disorders including major depression(Bahrami et al. 2020). While robustness checks found little evidence of pleiotropy, these methods rely on assumptions. Moreover, MR-Egger is known to give imprecise estimates(Burgess and Thompson 2017), and confidence intervals from MR-Egger models were wide. Thus, pleiotropy cannot be ruled out. The Mendelian randomization methods employed here assume any causal impact of BMI is linear – that a kg/m^2^ increase in BMI will have the same impact regardless of the child’s initial BMI. There is substantial evidence for a ‘J-shaped’ phenotypic association of BMI with common mental disorders, consistent with an impact of both high and low BMI on risk of depression or anxiety(McCrea, Berger, and King 2012; Gaysina et al. 2011; Geoffroy, Li, and Power 2014). Genetic methods exist for exposures with nonlinear effects but require much larger samples(Sun et al. 2019). If there exist nonlinear effects of BMI on mental health, rather than vice versa, our results may underestimate the effects of high BMI. Finally, the effects of BMI on emotional and behavioural functioning likely differ by age, and relationships may be substantially different for older children or adolescents. In particular, depressive symptoms do not tend to occur until the teenage years(Kwong et al. 2019) and observational associations of BMI and ADHD become clearer with age(Nigg et al. 2016). Work in larger samples of related individuals will be needed to precisely estimate the influence of a child’s BMI on their emotional and behavioural outcomes. MoBa is currently the largest individual study in which this approach can be applied, but new data are becoming available which will allow analyses of this kind within and across studies, such as through the Within Family Consortium https://www.withinfamilyconsortium.com/home/. Meanwhile, studies with extensive intergenerational information will be needed to fully explore mechanisms linking child outcomes to maternal BMI.

## Conclusion

Our results suggest that genetic variation associated with BMI in adulthood affects a child’s depressive and ADHD symptoms, but genetic variation associated with recalled childhood body size does not substantially affect these outcomes. There was little evidence that BMI affects anxiety. However, our estimates were imprecise, and these differences may be due to estimation error. There was little evidence that parental BMI affects a child’s ADHD or anxiety symptoms, but factors associated with maternal BMI may independently influence a child’s depressive symptoms. Genetic studies using unrelated individuals, or polygenic scores for adult BMI, may have overestimated the causal effects of a child’s own BMI.

## Data Availability

The consent given by the participants does not open for storage of data on an individual level in repositories or journals. Researchers who want access to data sets for replication should submit an application to. Access to data sets requires approval from The Regional Committee for Medical and Health Research Ethics in Norway and an agreement with MoBa.

## Acknowledgments

The Norwegian Mother, Father and Child Cohort Study is supported by the Norwegian Ministry of Health and Care Services and the Ministry of Education and Research. We are grateful to all the participating families in Norway who take part in this on-going cohort study. We thank the Norwegian Institute of Public Health (NIPH) for generating high-quality genomic data. This work was performed on the TSD (Tjeneste for Sensitive Data) facilities, owned by the University of Oslo, operated and developed by the TSD service group at the University of Oslo, IT-Department (USIT). The analyses were performed on resources provided by Sigma2 - the National Infrastructure for High Performance Computing and Data Storage in Norway. This research is part of the HARVEST collaboration, supported by the Research Council of Norway (#229624). We also thank the NORMENT Centre for providing genotype data, funded by the Research Council of Norway (#223273) and deCODE Genetics, South East Norway Health Authority and KG Jebsen Stiftelsen. We further thank the Center for Diabetes Research, the University of Bergen for providing genotype data and performing quality control and imputation of the data funded by the ERC AdG project SELECTionPREDISPOSED, Stiftelsen Kristian Gerhard Jebsen, Trond Mohn Foundation, the Research Council of Norway, the Novo Nordisk Foundation, the University of Bergen, and the Western Norway Health Authorities (Helse Vest).

## Data and code availability

The consent given by the participants does not open for storage of data on an individual level in repositories or journals. Researchers who want access to data sets for replication should submit an application to. Access to data sets requires approval from The Regional Committee for Medical and Health Research Ethics in Norway and an agreement with MoBa. Code used to generate these results is available at https://github.com/ammhughes/BMI-depressive-anxiety-and-ADHD-symptoms-in-MoBa

## Legend for Supplementary File 1

Supplementary File 1a. SNPs used in the polygenic score for adult BMI

Supplementary File 1b. SNPs used in the polygenic score for childhood body size BMI

Supplementary File 1c. SNPs used in the polygenic score for depression

Supplementary File 1d. SNPs used in the polygenic score for ADHD

## Figure titles and captions

## References

This includes references cited in Appendix 1 (Corfield et al. 2022; Auton et al. 2015; Kumar and Steer 2003; Choi and O’Reilly 2019)

Arafat, Subhi, and Camelia C. Minica. 2018. “Fetal Origins of Mental Disorders? An Answer Based on Mendelian Randomization.” Twin Research and Human Genetics. https://doi.org/10.1017/thg.2018.65.

Auton, Adam, Gonçalo R Abecasis, David M Altshuler, Richard M Durbin, Gonçalo R Abecasis, David R Bentley, Aravinda Chakravarti, et al. 2015. “A Global Reference for Human Genetic Variation.” Nature 526 (7571): 68–74. https://doi.org/10.1038/nature15393.

Bahrami, Shahram, Nils Eiel Steen, Alexey Shadrin, Kevin O’Connell, Oleksandr Frei, Francesco Bettella, Katrine v Wirgenes, et al. 2020. “Shared Genetic Loci Between Body Mass Index and Major Psychiatric Disorders: A Genome-Wide Association Study.” JAMA Psychiatry 77 (5): 503–12. https://doi.org/10.1001/jamapsychiatry.2019.4188.

Birmaher, Boris, David A. Brent, Laurel Chiappetta, Jeffrey Bridge, Suneeta Monga, and Marianne Baugher. 1999. “Psychometric Properties of the Screen for Child Anxiety Related Emotional Disorders (SCARED): A Replication Study.” Journal of the American Academy of Child and Adolescent Psychiatry. https://doi.org/10.1097/00004583-199910000-00011.

Blaine, Bruce. 2008. “Does Depression Cause Obesity?” Journal of Health Psychology. https://doi.org/10.1177/1359105308095977.

Bozoyan, Christiane, and Tobias Wolbring. 2018. “The Weight Wage Penalty: A Mechanism Approach to Discrimination.” European Sociological Review 34 (3): 254–67. https://doi.org/10.1093/esr/jcy009.

Brandkvist, Maria, Johan Håkon Bjørngaard, Rønnaug Astri ødegård, Bjørn Olav Åsvold, George Davey Smith, Ben Brumpton, Kristian Hveem, Tom G Richardson, and Gunnhild Åberge Vie. 2020. “Separating the Genetics of Childhood and Adult Obesity: A Validation Study of Genetic Scores for Body Mass Index in Adolescence and Adulthood in the HUNT Study.” Human Molecular Genetics 29 (24): 3966–73. https://doi.org/10.1093/hmg/ddaa256.

Brumpton, Ben, Eleanor Sanderson, Karl Heilbron, Fernando Pires Hartwig, Sean Harrison, Gunnhild Åberge Vie, Yoonsu Cho, et al. 2020. “Avoiding Dynastic, Assortative Mating, and Population Stratification Biases in Mendelian Randomization through within-Family Analyses.” Nature Communications. https://doi.org/10.1038/s41467-020-17117-4.

Burgess, Stephen, and Simon G Thompson. 2017. “Interpreting Findings from Mendelian Randomization Using the MR-Egger Method.” European Journal of Epidemiology 32 (5): 377–89. https://doi.org/10.1007/s10654-017-0255-x.

Chen, Qi, Arvid Sjölander, Niklas Långström, Alina Rodriguez, Eva Serlachius, Brian M D’Onofrio, Paul Lichtenstein, and Henrik Larsson. 2014. “Maternal Pre-Pregnancy Body Mass Index and Offspring Attention Deficit Hyperactivity Disorder: A Population-Based Cohort Study Using a Sibling-Comparison Design.” International Journal of Epidemiology 43 (1): 83–90. https://doi.org/10.1093/ije/dyt152.

Choi, Shing Wan, and Paul F O’Reilly. 2019. “PRSice-2: Polygenic Risk Score Software for Biobank-Scale Data.” GigaScience 8 (7). https://doi.org/10.1093/gigascience/giz082.

Corfield, Elizabeth C, Oleksandr Frei, Alexey A Shadrin, Zillur Rahman, Aihua Lin, Lavinia Athanasiu, Bayram Cevdet Akdeniz, et al. 2022. “The Norwegian Mother, Father, and Child Cohort Study (MoBa) Genotyping Data Resource: MoBaPsychGen Pipeline v.1.” BioRxiv, January, 2022.06.23.496289. https://doi.org/10.1101/2022.06.23.496289.

Cortese, Samuele, and Luca Tessari. 2017. “Attention-Deficit/Hyperactivity Disorder (ADHD) and Obesity: Update 2016.” Current Psychiatry Reports. https://doi.org/10.1007/s11920-017-0754-1.

Davies, Neil M., Michael v. Holmes, and George Davey Smith. 2018. “Reading Mendelian Randomisation Studies: A Guide, Glossary, and Checklist for Clinicians.” BMJ (Online). https://doi.org/10.1136/bmj.k601.

Davies, Neil M, Laurence J Howe, Ben Brumpton, Alexandra Havdahl, David M Evans, and George Davey Smith. 2019. “Within Family Mendelian Randomization Studies.” Human Molecular Genetics 28 (R2): R170–79. https://doi.org/10.1093/hmg/ddz204.

Demontis, Ditte, Raymond K. Walters, Joanna Martin, Manuel Mattheisen, Thomas D. Als, Esben Agerbo, Gísli Baldursson, et al. 2019. “Discovery of the First Genome-Wide Significant Risk Loci for Attention Deficit/Hyperactivity Disorder.” Nature Genetics. https://doi.org/10.1038/s41588-018-0269-7.

Edlow, Andrea G. 2017. “Maternal Obesity and Neurodevelopmental and Psychiatric Disorders in Offspring.” Prenatal Diagnosis 37 (1): 95–110. https://doi.org/ https://doi.org/10.1002/pd.4932.

Garrido-Miguel, Miriam, Iván Cavero-Redondo, Celia Álvarez-Bueno, Fernando Rodríguez-Artalejo, Luis A. Moreno, Jonatan R. Ruiz, Wolfgang Ahrens, and Vicente Martínez-Vizcaíno. 2019. “Prevalence and Trends of Overweight and Obesity in European Children from 1999 to 2016: A Systematic Review and Meta-Analysis.” JAMA Pediatrics. https://doi.org/10.1001/jamapediatrics.2019.2430.

Gaysina, D., M. Hotopf, M. Richards, I. Colman, D. Kuh, and R. Hardy. 2011. “Symptoms of Depression and Anxiety, and Change in Body Mass Index from Adolescence to Adulthood: Results from a British Birth Cohort.” Psychological Medicine. https://doi.org/10.1017/S0033291710000346.

Geoffroy, M.-C., L. Li, and C. Power. 2014. “Depressive Symptoms and Body Mass Index: Co-Morbidity and Direction of Association in a British Birth Cohort Followed over 50 Years.” Psychological Medicine 44 (12): 2641–52. https://doi.org/10.1017/S0033291714000142.

Geuijen, Pauline M., Jan K. Buitelaar, Ellen A. Fliers, Athanasios Maras, Lizanne J.S. Schweren, Jaap Oosterlaan, Pieter J. Hoekstra, Barbara Franke, Catharina A. Hartman, and Nanda N. Rommelse. 2019. “Overweight in Family Members of Probands with ADHD.” European Child and Adolescent Psychiatry. https://doi.org/10.1007/s00787-019-01331-7.

Giel, Katrin Elisabeth, Ansgar Thiel, Martin Teufel, Jochen Mayer, and Stephan Zipfel. 2010. “Weight Bias in Work Settings - A Qualitative Review.” Obesity Facts 3: 33–40. https://doi.org/10.1159/000276992.

Glavin, Kari, Mathieu Roelants, Bjørn Heine Strand, Pétur B Júlíusson, Kari Kveim Lie, Sølvi Helseth, and Ragnhild Hovengen. 2014. “Important Periods of Weight Development in Childhood: A Population-Based Longitudinal Study.” BMC Public Health 14 (February): 160. https://doi.org/10.1186/1471-2458-14-160.

Griffiths, Lucy J, Carol Dezateux, and Andrew Hill. 2011. “Is Obesity Associated with Emotional and Behavioural Problems in Children? Findings from the Millennium Cohort Study.” International Journal of Pediatric Obesity : IJPO : An Official Journal of the International Association for the Study of Obesity 6 (2–2): e423–32. https://doi.org/10.3109/17477166.2010.526221.

Hemani, Gibran, Jack Bowden, and George Davey Smith. 2018. “Evaluating the Potential Role of Pleiotropy in Mendelian Randomization Studies.” Human Molecular Genetics. https://doi.org/10.1093/hmg/ddy163.

Hemani, Gibran, Jie Zheng, Benjamin Elsworth, Kaitlin H. Wade, Valeriia Haberland, Denis Baird, Charles Laurin, et al. 2018. “The MR-Base Platform Supports Systematic Causal Inference across the Human Phenome.” ELife. https://doi.org/10.7554/eLife.34408.

Hesbacher, P T, K Rickels, R J Morris, H Newman, and H Rosenfeld. 1980. “Psychiatric Illness in Family Practice.” The Journal of Clinical Psychiatry 41 (1): 6–10.

Hope, Steven, Nadia Micali, Jessica Deighton, and Catherine Law. 2019. “Maternal Mental Health at 5 Years and Childhood Overweight or Obesity at 11 Years: Evidence from the UK Millennium Cohort Study.” International Journal of Obesity. https://doi.org/10.1038/s41366-018-0252-5.

Hope, Steven, Anna Pearce, Catherine Chittleborough, Jessica Deighton, Amelia Maika, Nadia Micali, Murthy Mittinty, Catherine Law, and John Lynch. 2019. “Temporal Effects of Maternal Psychological Distress on Child Mental Health Problems at Ages 3, 5, 7 and 11: Analysis from the UK Millennium Cohort Study.” Psychological Medicine. https://doi.org/10.1017/S0033291718001368.

Howe, Laura D, Roshni Kanayalal, Sean Harrison, Robin N Beaumont, Alisha R Davies, Timothy M Frayling, Neil M Davies, et al. 2019. “Effects of Body Mass Index on Relationship Status, Social Contact and Socio-Economic Position: Mendelian Randomization and within-Sibling Study in UK Biobank.” International Journal of Epidemiology. https://doi.org/10.1093/ije/dyz240.

Howe, Laurence J, Michel G Nivard, Tim T Morris, Ailin F Hansen, Humaira Rasheed, Yoonsu Cho, Geetha Chittoor, et al. 2022. “Within-Sibship Genome-Wide Association Analyses Decrease Bias in Estimates of Direct Genetic Effects.” Nature Genetics 54 (5): 581–92. https://doi.org/10.1038/s41588-022-01062-7.

Kessler, Ronald C, Lenard Adler, Minnie Ames, Olga Demler, Steve Faraone, Eva Hiripi, Mary J Howes, et al. 2005. “The World Health Organization Adult ADHD Self-Report Scale (ASRS): A Short Screening Scale for Use in the General Population.” Psychological Medicine 35 (2): 245–56. https://doi.org/10.1017/s0033291704002892.

Kong, Augustine, Gudmar Thorleifsson, Michael L. Frigge, Bjarni J. Vilhjalmsson, Alexander I. Young, Thorgeir E. Thorgeirsson, Stefania Benonisdottir, et al. 2018. “The Nature of Nurture: Effects of Parental Genotypes.” Science. https://doi.org/10.1126/science.aan6877.

Kovess-Masfety, Viviane, Mathilde M Husky, Katherine Keyes, Ava Hamilton, Ondine Pez, Adina Bitfoi, Mauro Giovanni Carta, et al. 2016. “Comparing the Prevalence of Mental Health Problems in Children 6-11 across Europe.” Social Psychiatry and Psychiatric Epidemiology 51 (8): 1093–1103. https://doi.org/10.1007/s00127-016-1253-0.

Kumar, Geetha, and Robert A Steer. 2003. “Factorial Validity of the Conners’ Parent Rating Scale-Revised: Short Form with Psychiatric Outpatients.” Journal of Personality Assessment 80 (3): 252–59. https://doi.org/10.1207/S15327752JPA8003_04.

Kwong, Alex S F, David Manley, Nicholas J Timpson, Rebecca M Pearson, Jon Heron, Hannah Sallis, Evie Stergiakouli, Oliver S P Davis, and George Leckie. 2019. “Identifying Critical Points of Trajectories of Depressive Symptoms from Childhood to Young Adulthood.” Journal of Youth and Adolescence 48 (4): 815–27. https://doi.org/10.1007/s10964-018-0976-5.

Li, Lin, Tyra Lagerberg, Zheng Chang, Samuele Cortese, Mina A Rosenqvist, Catarina Almqvist, Brian M D’Onofrio, et al. 2020. “Maternal Pre-Pregnancy Overweight/Obesity and the Risk of Attention-Deficit/Hyperactivity Disorder in Offspring: A Systematic Review, Meta-Analysis and Quasi-Experimental Family-Based Study.” International Journal of Epidemiology 31: 1–19. https://doi.org/10.1093/ije/dyaa040.

Lindberg, Louise, Emilia Hagman, Pernilla Danielsson, Claude Marcus, and Martina Persson. 2020. “Anxiety and Depression in Children and Adolescents with Obesity: A Nationwide Study in Sweden.” BMC Medicine. https://doi.org/10.1186/s12916-020-1498-z.

Magnus, Per, Charlotte Birke, Kristine Vejrup, Anita Haugan, Elin Alsaker, Anne Kjersti Daltveit, Marte Handal, et al. 2016. “Cohort Profile Update: The Norwegian Mother and Child Cohort Study (MoBa).” International Journal of Epidemiology 45 (2): 382–88. https://doi.org/10.1093/ije/dyw029.

Manichaikul, Ani, Josyf C. Mychaleckyj, Stephen S. Rich, Kathy Daly, Michèle Sale, and Wei Min Chen. 2010. “Robust Relationship Inference in Genome-Wide Association Studies.” Bioinformatics 26 (22): 2867–73. https://doi.org/10.1093/bioinformatics/btq559.

Martins-Silva, Thais, Juliana dos Santos Vaz, Mara Helena Hutz, Angélica Salatino-Oliveira, Júlia Pasqualini Genro, Fernando Pires Hartwig, Carlos Renato Moreira-Maia, Luis Augusto Rohde, Maria Carolina Borges, and Luciana Tovo-Rodrigues. 2019. “Assessing Causality in the Association between Attention-Deficit/Hyperactivity Disorder and Obesity: A Mendelian Randomization Study.” International Journal of Obesity. https://doi.org/10.1038/s41366-019-0346-8.

McCrea, R. L., Y. G. Berger, and M. B. King. 2012. “Body Mass Index and Common Mental Disorders: Exploring the Shape of the Association and Its Moderation by Age, Gender and Education.” International Journal of Obesity. https://doi.org/10.1038/ijo.2011.65.

Millard, Louise A.C., Neil M. Davies, Kate Tilling, Tom R. Gaunt, and George Davey Smith. 2019. “Searching for the Causal Effects of Body Mass Index in over 300 000 Participants in UK Biobank, Using Mendelian Randomization.” PLoS Genetics. https://doi.org/10.1371/journal.pgen.1007951.

Morris, Tim T., Neil M. Davies, Gibran Hemani, and George Davey Smith. 2020. “Population Phenomena Inflate Genetic Associations of Complex Social Traits.” Science Advances. https://doi.org/10.1126/sciadv.aay0328.

Nigg, Joel T, Jeanette M Johnstone, Erica D Musser, Hilary Galloway Long, Michael T Willoughby, and Jackilen Shannon. 2016. “Attention-Deficit/Hyperactivity Disorder (ADHD) and Being Overweight/Obesity: New Data and Meta-Analysis.” Clinical Psychology Review 43 (February): 67–79. https://doi.org/10.1016/j.cpr.2015.11.005.

Patalay, Praveetha, and Charlotte A Hardman. 2019. “Comorbidity, Codevelopment, and Temporal Associations Between Body Mass Index and Internalizing Symptoms From Early Childhood to Adolescence.” JAMA Psychiatry 76 (7): 721–29. https://doi.org/10.1001/jamapsychiatry.2019.0169.

Puhl, Rebecca M., Melanie M. Wall, Chen Chen, S. Bryn Austin, Marla E. Eisenberg, and Dianne Neumark-Sztainer. 2017. “Experiences of Weight Teasing in Adolescence and Weight-Related Outcomes in Adulthood: A 15-Year Longitudinal Study.” Preventive Medicine. https://doi.org/10.1016/j.ypmed.2017.04.023.

Quek, Ying-Hui, Wilson W S Tam, Melvyn W B Zhang, and Roger C M Ho. 2017. “Exploring the Association between Childhood and Adolescent Obesity and Depression: A Meta-Analysis.” Obesity Reviews : An Official Journal of the International Association for the Study of Obesity 18 (7): 742–54. https://doi.org/10.1111/obr.12535.

Richardson, Tom G, Eleanor Sanderson, Benjamin Elsworth, Kate Tilling, and George Davey Smith. 2020. “Use of Genetic Variation to Separate the Effects of Early and Later Life Adiposity on Disease Risk: Mendelian Randomisation Study.” BMJ 369 (May): m1203. https://doi.org/10.1136/bmj.m1203.

Rubino, Francesco, Rebecca M. Puhl, David E. Cummings, Robert H. Eckel, Donna H. Ryan, Jeffrey I. Mechanick, Joe Nadglowski, et al. 2020. “Joint International Consensus Statement for Ending Stigma of Obesity.” Nature Medicine 26 (4): 485–97. https://doi.org/10.1038/s41591-020-0803-x.

Russell, Abigail Emma, Tamsin Ford, Rebecca Williams, and Ginny Russell. 2016. “The Association Between Socioeconomic Disadvantage and Attention Deficit/Hyperactivity Disorder (ADHD): A Systematic Review.” Child Psychiatry and Human Development. https://doi.org/10.1007/s10578-015-0578-3.

Sadler, Katharine, Tim Vizard, Tamsin Ford, Anna Goodman, Robert Goodman, and Sally McManus. 2018. “Mental Health of Children and Young People in England, 2017.” https://digital.nhs.uk/data-and-information/publications/statistical/mental-health-of-children-and-young-people-in-england/2017/2017.

Sanchez, C.E., A. Sabhlok, K. Russell, A. Majors, S.H. Kollins, and B.F. Fuemmeler. 2018. “Maternal Pre-Pregnancy Obesity and Child Neurodevelopmental Outcomes: A Meta-Analysis.” Obesity Reviews. https://doi.org/10.1111/obr.12643.

Sayal, Kapil, Vibhore Prasad, David Daley, Tamsin Ford, and David Coghill. 2018. “ADHD in Children and Young People: Prevalence, Care Pathways, and Service Provision.” The Lancet Psychiatry 5 (2): 175–86. https://doi.org/ https://doi.org/10.1016/S2215-0366(17)30167-0.

Silva, Raul R., Murray Alpert, Enrique Pouget, Victoria Silva, Sarah Trosper, Kimberly Reyes, and Steven Dummit. 2005. “A Rating Scale for Disruptive Behavior Disorders, Based on the DSM-IV Item Pool.” Psychiatric Quarterly. https://doi.org/10.1007/s11126-005-4966-x.

Silventoinen, Karri, Aline Jelenkovic, Reijo Sund, Yoon-Mi Hur, Yoshie Yokoyama, Chika Honda, Jacob vB Hjelmborg, et al. 2016. “Genetic and Environmental Effects on Body Mass Index from Infancy to the Onset of Adulthood: An Individual-Based Pooled Analysis of 45 Twin Cohorts Participating in the COllaborative Project of Development of Anthropometrical Measures in Twins (CODATwins).” The American Journal of Clinical Nutrition 104 (2): 371–79. https://doi.org/10.3945/ajcn.116.130252.

Spiller, Wes, Neil M. Davies, and Tom M. Palmer. 2019. “Software Application Profile: Mrrobust -A Tool for Performing Two-Sample Summary Mendelian Randomization Analyses.” International Journal of Epidemiology. https://doi.org/10.1093/ije/dyy195.

Sun, Yi-Qian, Stephen Burgess, James R Staley, Angela M Wood, Steven Bell, Stephen K Kaptoge, Qi Guo, et al. 2019. “Body Mass Index and All Cause Mortality in HUNT and UK Biobank Studies: Linear and Non-Linear Mendelian Randomisation Analyses.” BMJ 364 (March): l1042. https://doi.org/10.1136/bmj.l1042.

Thompson, Melissa, Lindsey Wilkinson, and Hyeyoung Woo. 2020. “Social Characteristics as Predictors of ADHD Labeling across the Life Course.” Society and Mental Health, May, 2156869320916535. https://doi.org/10.1177/2156869320916535.

Tyrrell, Jessica, Anwar Mulugeta, Andrew R. Wood, Ang Zhou, Robin N. Beaumont, Marcus A. Tuke, Samuel E. Jones, et al. 2019. “Using Genetics to Understand the Causal Influence of Higher BMI on Depression.” International Journal of Epidemiology. https://doi.org/10.1093/ije/dyy223.

Vizard, Tim, Katharine Sadler, Tamsin Ford, Tamsin Newlove-Delgado, Sally McManus, Jodie Marcheselli, Franziska Davis, Tracy Williams, Charlotte Leach, Dhriti Mandalia, and Cher Cartwright. 2020. “Mental Health of Children and Young People in England, 2020: Wave 1 Follow up to the 2017 Survey.” https://files.digital.nhs.uk/AF/AECD6B/mhcyp_2020_rep_v2.pdf.

Vogel, Suzan W N, Denise Bijlenga, Marjolein Tanke, Tannetje I Bron, Kristiaan B van der Heijden, Hanna Swaab, Aartjan T F Beekman, and J J Sandra Kooij. 2015. “Circadian Rhythm Disruption as a Link between Attention-Deficit/Hyperactivity Disorder and Obesity?” Journal of Psychosomatic Research 79 (5): 443–50. https://doi.org/ https://doi.org/10.1016/j.jpsychores.2015.10.002.

Walter, S., M. M. Glymour, K. Koenen, L. Liang, E. J. Tchetgen Tchetgen, M. Cornelis, S. C. Chang, et al. 2015. “Do Genetic Risk Scores for Body Mass Index Predict Risk of Phobic Anxiety? Evidence for a Shared Genetic Risk Factor.” Psychological Medicine. https://doi.org/10.1017/S0033291714001226.

Wray, Naomi R., Stephan Ripke, Manuel Mattheisen, MacIej Trzaskowski, Enda M. Byrne, Abdel Abdellaoui, Mark J. Adams, et al. 2018. “Genome-Wide Association Analyses Identify 44 Risk Variants and Refine the Genetic Architecture of Major Depression.” Nature Genetics. https://doi.org/10.1038/s41588-018-0090-3.

Yengo, Loic, Julia Sidorenko, Kathryn E. Kemper, Zhili Zheng, Andrew R. Wood, Michael N. Weedon, Timothy M. Frayling, et al. 2018. “Meta-Analysis of Genome-Wide Association Studies for Height and Body Mass Index in ∼700000 Individuals of European Ancestry.” Human Molecular Genetics 27 (20): 3641–49. https://doi.org/10.1093/hmg/ddy271.

