## Appendix 1 for "Body mass index and childhood symptoms of depression, anxiety, and attention-deficit hyperactivity disorder: a within-family Mendelian randomization study"

### Appendix 1: additional methods, figures, and small tables

*Contents of this file:*

Additional Methods:


1. MoBa study details
2. Genotyping and imputation
3. Genetic quality control
4. Multiple imputation of phenotypes
5. Polygenic score construction
6. Model equations


Additional Results:


1. Comparison of analytic sample and excluded participants


Additional Figures:


Appendix 1 – Figure 1. Flow chart of inclusion and exclusion of MoBa participants into the study sample
Appendix 1 – Figure 2. Association of child's BMI polygenic scores with principal components of ancestry
Appendix 1 – Figure 3. Associations of child's BMI polygenic scores with ancestry, adjusted for parental polygenic scores.


Additional Tables:


Appendix 1 – Table 1. Descriptive statistics of full MoBa sample
Appendix 1 – Table 2. Descriptive statistics of analytic sample, unimputed data
Appendix 1 – Table 3. BMI and symptoms of depression, anxiety, and ADHD at age 8, adult BMI PGS
Appendix 1 – Table 4. BMI and symptoms of depression, anxiety, and ADHD at age 8, childhood body size PGS
Appendix 1 – Table 5. Associations of polygenic scores within parental pairs
Appendix 1 – Table 6. BMI and symptoms of depression, anxiety, and ADHD at age 8, adult BMI PGS, log-transformed outcomes
Appendix 1 – Table 7. BMI and symptoms of depression, anxiety, and ADHD at age 8, childhood body size PGS, log-transformed outcomes
Appendix 1 – Table 8. Robustness checks for classic MR models
Appendix 1 – Table 9. Robustness checks for within-family MR (within-family MR) models
Appendix 1 – Table 10. BMI and symptoms of depression, anxiety, and ADHD at age 8 in MoBa, adult BMI PGS, complete-case analysis
Appendix 1 – Table 11. BMI and symptoms of depression, anxiety, and ADHD at age 8 in MoBa, childhood body size PGS, complete-case analysis
Appendix 1 – Table 12. Non-genetic associations of BMI quintiles with symptoms of depression, anxiety, and ADHD at age 8 in MoBa
Appendix 1 – Table 13. BMI and symptoms of depression, anxiety, and ADHD at age 8, adult BMI PGS, adjusted for parental education
Appendix 1 – Table 14. BMI and symptoms of depression, anxiety, and ADHD at age 8, childhood body size PGS, adjusted for parental education


*The following large tables are provided in a separate excel file (Supplementary File 1):*


Supplementary File 1a. SNPs used in the polygenic score for adult BMI
Supplementary File 1b. SNPs used in the polygenic score for childhood body size

Supplementary File 1c. SNPs used in the polygenic score for depression
Supplementary File 1d. SNPs used in the polygenic score for ADHD

**Additional Methods**

1. MoBa study details

This study is based on the Norwegian Mother, Father and Child Cohort Study (MoBa) and uses data from the Medical Birth Registry of Norway (MBRN). The Medical Birth Registry (MBRN) is a national health registry containing information about all births in Norway. The current analysis is based on version 12 of the quality-assured data files released for research in January 2019. The establishment of MoBa and initial data collection was based on a license from the Norwegian Data Protection Agency and approval from The Regional Committees for Medical and Health Research Ethics. The MoBa cohort is now based on regulations related to the Norwegian Health Registry Act. The current study was approved by The Regional Committees for Medical and Health Research Ethics (2016/1702).

The Norwegian Mother, Father and Child Cohort Study is supported by the Norwegian Ministry of Health and Care Services and the Ministry of Education and Research. We are grateful to all the participating families in Norway who take part in this on-going cohort study.

2. Genotyping

Genotyping of MoBa has been conducted through multiple research projects, spanning several years. The research projects (HARVEST, SELECTIONpreDISPOSED, and NORMENT) provided genotype data to MoBa Genetics. In total, 238,001 MoBa samples were sent to be genotyped in 24 genotyping batches. This was carried out at 3 centres (1. Genomics Core Facility, Trondheim, Norway, 2. ERASMUS MC, Rotterdam, Netherlands, and 3. deCODE Genetics, Reykjavik, Iceland) using six genotyping arrays. The 24 batches had varying selection criteria; this included a batch of ADHD child cases and their parents, and another of matched control children and their parents. Detailed information on batch selection and the genotyping process are described elsewhere (Corfield et al. 2022).

3. Genetic quality control

Pre-imputation QC, phasing, imputation, and post-imputation QC were carried out according to the MoBaPsychGen pipeline, which includes QC on both single nucleotide polymorphism (SNP) and individual level, and whose full details are provided elsewhere (Corfield et al. 2022). Output of the pipeline included 207,569 unique individuals and 6,981,748 SNPs after post-imputation QC; unique individuals comprised 76,577 children, 53,358 fathers, and 77,634 mothers. Phasing and imputation were performed using the publicly available Haplotype Reference Consortium release 1.1 panel as a reference. Information from the Medical Birth Registry of Norway and MoBa questionnaires were used to identify biological sex, year of birth, reported parent-offspring (PO) relationships, and, in the offspring generation, multiple births. Ancestry outliers were identified based on principal component analysis with the 1000 Genomes phase 1 unrelated data (1,083 individuals)(Auton et al. 2015). Approximately 95% of the participants were identified as having European ancestry. Ancestry outliers were removed based on visual inspection, using pairwise plots for the first seven principal components. Similarly, principal component analysis was used to identify substructure within each subpopulation of all MoBa batches. PCs were first estimated in founders only, and non-founders projected into the PC space of the founders. Outliers were removed based on visual inspection, using pairwise plots for the first ten principal components.

Relatedness was inferred using KING version 2.2.5(Manichaikul et al. 2010). In the presence of admixture, KING accurately infers MZ twin or duplicate pairs (kinship coefficient > 0.3540), first-degree (PO, FS, DZ twin pairs; kinship coefficient range 0.1770 - 0.3540), second-degree (HS, GO, AUNN; kinship coefficient range 0.0884 – 0.1770), and third-degree (first cousins; kinship coefficient range 0.0442 – 0.0884) relationships. After within-family and between-family relationships were confirmed by genetic data, a check for Mendelian errors (ME) was performed in PLINK. The ME check included families with one or two parents present in the data. Families with > 5% errors and SNPs with > 1% errors were removed. The remaining ME were set to missing.

**4.** Multiple imputation

Multiple imputation by chained equations was performed in STATAv16 to estimate missing phenotypic information for the 40,949 trios with complete genetic data. 100 imputed datasets were produced and analysis across these datasets conducted with STATA's mi estimate commands. The imputation model included all BMI variables used in the main analyses (child's BMI at age 8, mother's pre-pregnancy BMI and father's BMI as reported at 17 weeks gestation), the child's sex and year of birth, and other phenotypic covariates used in non-genetic models, including mother's and father's smoking status reported at 17 weeks gestation, mother's and father's depressive/anxiety symptoms (using selected items from the 25-item Hopkins Checklist(Hesbacher et al. 1980)), and ADHD symptoms (from the 6-item adult ADHD self-report scale(Kessler et al. 2005)), maternal parity at the child's birth, and family socioeconomic characteristics, including parental educational qualifications and categorical variables of income and subjective financial strain. Variables from the birth registry file were also included as auxiliary variables: the mother's marital status, the age of the mother and father, and the child's birthweight and length. Approximately normally-distributed continuous variables including BMI were imputed using truncated regression, specifying as upper and lower limits the smallest and largest values observed in the full MoBa sample. Ordered categorical variables were imputed with ordered logistic regression. There was no missingness in genetic information within the analytic sample. Polygenic scores for adult BMI, childhood body size, depression, ADHD, and educational attainment were included on the right-hand side of the imputation equations, along with indicators for genotyping centre and chip and the 20 principal components of ancestry for all individuals. Continuous variables which were not normally-distributed were imputed with predictive mean matching, specifying knn(10). This included child's depressive and anxiety symptoms at age 8 (SMFQ and SCARED summary scores), mother's and father's depressive/anxiety symptoms at 17 weeks gestation (summary scores based on items from the 25-item Hopkins Checklist(Hesbacher et al. 1980), and mother's and father's ADHD symptoms from the 6-item adult ADHD self-report scale(Kessler et al. 2005). To facilitate analysis of ADHD inattention and hyperactivity subscales, the two subscales were imputed, again with predictive mean matching, and the full scale calculated post-imputation with mi passive. An earlier measure of the child's ADHD symptoms at 5yrs, based on questions from the Short-Form Conners Parent Rating Scale(Kumar and Steer 2003), was included as an auxiliary variable. The percentage of imputed data in the analytic sample for each variable was: mother's education 3.4%, father's education 2.2%, mother's smoking 2.0%, father's smoking 0.7%, mother's depressive symptoms 3.2%, father's depressive symptoms 7.1%, mother's ADHD symptoms 40.9%, father's ADHD symptoms 60.1%, mother's BMI: 4.0%, father's BMI: 3.8%, child's BMI at age 8: 60.6%, child's depressive symptoms at age 8: 54.2%, child's anxiety symptoms at age 8: 54.1%, child's ADHD symptoms (inattention and hyperactivity) both 54.1%.

### 5. Polygenic score construction

Beginning with the full GWAS results for each phenotype(Yengo et al. 2018; Richardson et al. 2020; Wray et al. 2018; Demontis et al. 2019), R was used to subset SNPs included in full GWAS results to SNPs also available in the quality controlled MoBa data. From SNPs available in both, independent genome-wide significant associations were identified by clumping in MRBase, specifying r=0.01 and $p<5.0x10^{-8}$ (Hemani et al. 2018). First subsetting to SNPs available in MoBa and then clumping within these avoids the need for an additional step identifying proxy SNPs. This left 954 SNPs associated with adult BMI, and 321 associated with childhood body size.

Dosage data for MoBa participants for the genetic variants relevant to each polygenic score were extracted from the final quality controlled -bfiles using PLINK, and .raw files imported to STATA. SNPs were harmonized by comparing effect alleles in the GWAS and reference alleles in MoBa.

Polygenic scores were calculated as the sum of the number of effect alleles (0,1, or 2) multiplied in each case by the harmonized SNP-coefficient from the GWAS. There was a very small amount of missingness in individual SNPs due to quality control measures. Among SNPs included in the adult BMI PGS, the median proportion of individuals missing the SNP was 0.001 and the maximum 0.04. For SNPs included in the childhood body size PGS, equivalent values were 0.002 and 0.04, for SNPs included in the depression and the ADHD PGS these were 0.003 and 0.01. Where individual SNP information was missing for an individual, the sample mean for the number of effect alleles (between 0 and 2) was added for that SNP.

### 6. Model equations

In within-family MR models, we used polygenic scores for all members of a child-mother-child trio to instrument the BMI of all three individuals. Within-families MR models were adjusted for child's sex, the first 20 principal components of ancestry for the child and both parents, and the genotyping chip and centre of the child and both parents.

In Stata, this was specified in the form:

ivregress 2sls outcome (c_bmi m_bmi f_bmi = c_pgs m_pgs f_pgs) sex c_PC* m_PC* f_PC* c_genotyping_centre* m_ genotyping_centre* f_ genotyping_centre* c_genotyping_chip* m_ genotyping_chip* f_ genotyping_chip*

This can be represented with the following equations:

The outcomes (depressive, anxiety, and ADHD symptoms) can be expressed as:

$$y_i = \beta_0 + \beta_1 X_{ic} + \beta_2 X_{im} + \beta_3 X_{if} + \beta_c C + e_i$$

The three exposures are offspring's, mother's, and father's BMI:

$$X_{ic} = \gamma_{oc} + \gamma_{1c} Z_{ic} + \gamma_{2c} Z_{im} + \gamma_{3c} Z_{if} + \gamma_{4c} C + v_{ic}$$

$$X_{im} = \gamma_{om} + \gamma_{1m} Z_{ic} + \gamma_{2m} Z_{im} + \gamma_{3m} Z_{if} + \gamma_{4m} C + v_{im}$$

$$X_{if} = \gamma_{oc} + \gamma_{1f} Z_{ic} + \gamma_{2f} Z_{im} + \gamma_{3f} Z_{if} + \gamma_{4f} C + v_{if}$$

where:

$y_i$=outcome
$X_{ic}$=child's BMI
$X_{im}$=mother's BMI
$X_{if}$=father's BMI
$Z_{ic}$=child's polygenic score
$Z_{im}$=mother's polygenic score
$Z_{if}$=father's polygenic score
$C$=covariates including principal components for offspring, mother and father and genotyping centre and chip
$e_i$= error term for the outcome equation
$v_{ic}$= error term for the child exposure (BMI) equation
$v_{im}$= error term for the mother exposure (BMI) equation
$v_{if}$= error term for the father exposure (BMI) equation

**Additional Results**

1. Comparison of analytic sample and excluded participants

To assess if participants included in the analytic sample (N=40,949) differed from others in the birth registry file (N=72,742) (Appendix 1 - Figure 1), we conducted t-tests and chi-squared tests for key characteristics at birth, BMI, and outcomes using unimputed data. Reflecting the large size of the sample population, several differences reached statistical significance. Children in the analytic sample did not differ from those excluded on sex but were slightly older (mean year of birth: 2005.4 vs 2004.5). They had slightly higher birthweight (mean=3.6kg vs 3.4kg) and slightly younger fathers (mean paternal 32.6 vs 32.8 years). Mothers and fathers included in the analytic sample had slightly higher educational qualifications (e.g., 24.1% and 24.3% of mothers and fathers respectively had 4 years of college education, against 21.0% and 21.5% for those not included). At the time of the child's birth, mothers in the analytic sample had fewer existing children (e.g., 46.8% vs 42.4% had none), were more likely to be married or cohabiting (97.4% vs 94.4%), and less likely to have smoked in pregnancy (e.g., daily smoking: 4.5% vs 6.3%). Fathers in the analytic sample were also more likely to have stopped smoking during the pregnancy (20.9% vs 15.8%). There was little of a difference in BMI for mothers, fathers, or children. Children in the analytic sample had slightly lower depressive symptoms (mean SMFQ=1.81 vs 1.91),anxiety symptoms (mean SCARED: 1.04 vs 1.00),and ADHD symptoms (mean RS-DBD ADHD: 8.4 vs 8.7). Descriptive characteristics of the full MoBa sample are in Appendix 1 - Table 1.

**Appendix 1 - Figure 1: Flow chart of inclusion and exclusion of MoBa participants into the study sample**

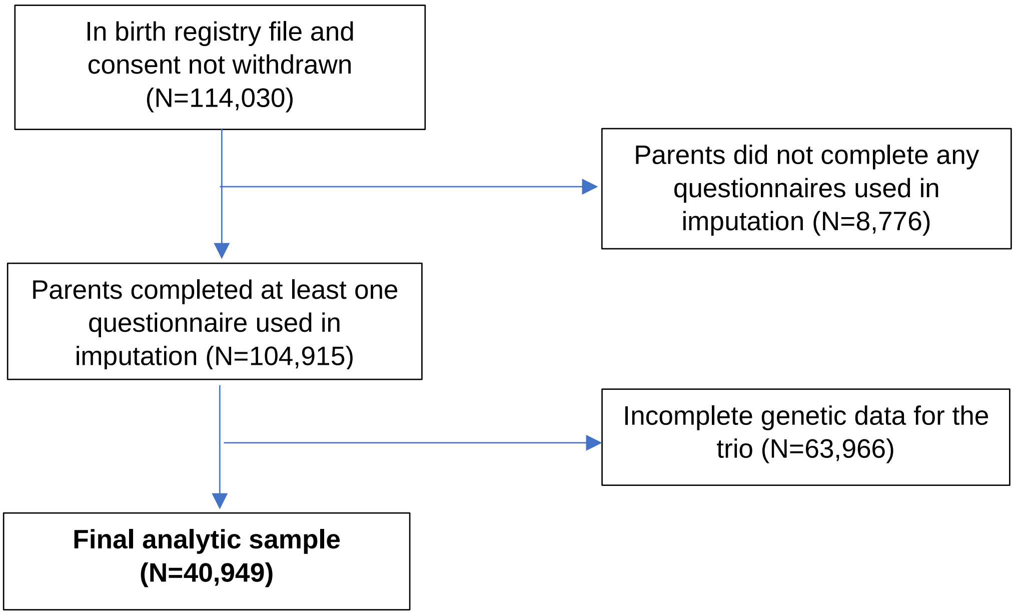

### Appendix 1 - Figure 2: Associations of child's BMI polygenic scores with ancestry

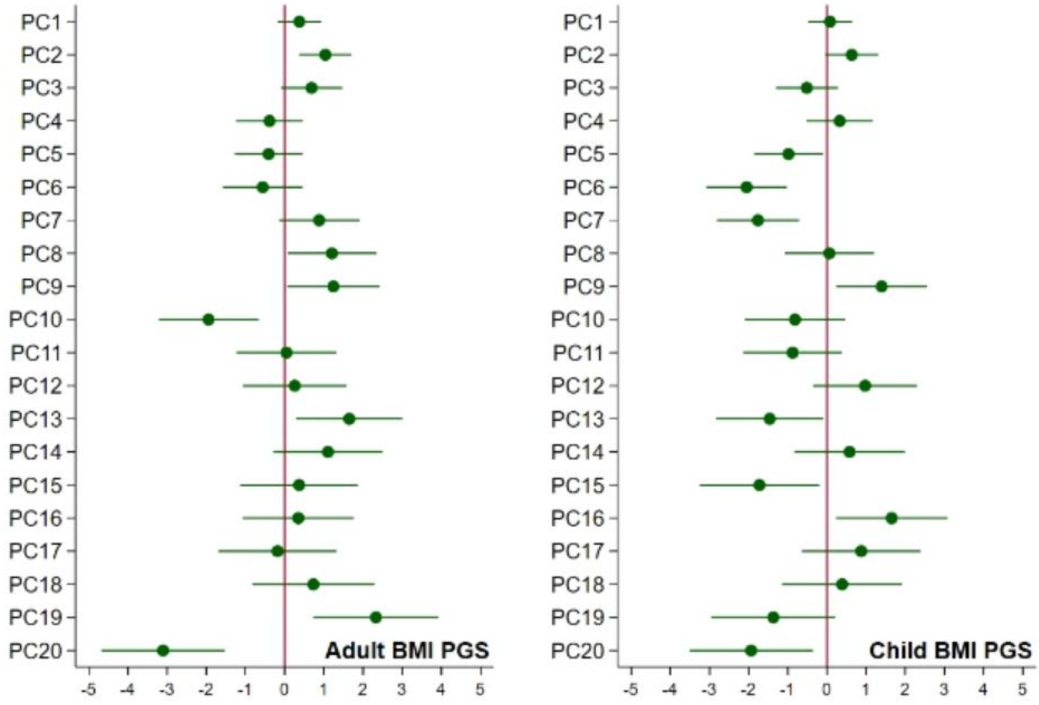


Caption: associations of the child's polygenic scores for BMI and the child's principal components of ancestry, adjusted for the child's genotyping centre and chip.

### Appendix 1 - Figure 3: Associations of child's BMI polygenic scores with ancestry, adjusted for parental polygenic scores.

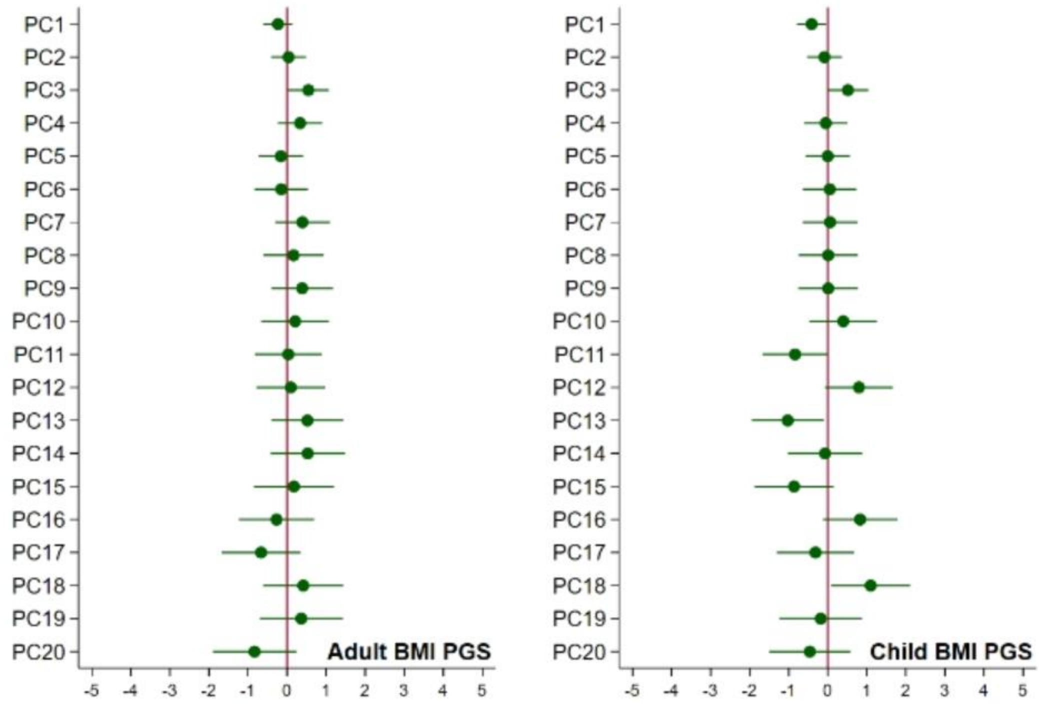


Caption: associations of the child's polygenic scores for BMI and the child's principal components of ancestry, adjusted for the child's genotyping centre and chip and the parents' polygenic scores.

**Appendix 1 - Table 1: Descriptive statistics of full MoBa sample (N=113,691)[a]**

| Continuous variables | | mean | SD | N obs |
|---|---|---|---|---|
| Maternal age at birth | | 30.1 | 4.6 | 113,691 |
| Paternal age at birth | | 32.7 | 5.5 | 113,172 |
| Maternal depressive/anxiety symptoms, based on 5 items from the Hopkins Symptoms Checklist-25 (SCL-25)[b] | | 1.3 | 2.0 | 100,570 |
| Paternal depressive/anxiety symptoms, based on 8 items from the Hopkins Symptoms Checklist-25 (SCL-25)[c] | | 1.2 | 2.1 | 77,018 |
| Maternal ADHD symptoms: adult ADHD self-report scale[d] | | 6.6 | 3.5 | 56,255 |
| Paternal ADHD symptoms: adult ADHD self-report scale[e] | | 8.2 | 3.2 | 34,425 |
| Maternal pre-pregnancy BMI (kg/m$^2$) | | 24.0 | 4.1 | 100,060 |
| Paternal BMI (kg/m$^2$) | | 25.9 | 3.3 | 79,536 |
| Child's BMI at age 8 (kg/m$^2$) | | 16.2 | 2.0 | 36,894 |
| Child depressive symptoms age 8: Short Mood and Feelings Questionnaire (SMFQ)[f] | | 1.9 | 2.4 | 43,065 |
| Child anxiety symptoms age 8: Screen for Child Anxiety Related Disorders (SCARED)[g] | | 1.0 | 1.2 | 43,298 |
| Child ADHD symptoms age 8: Parent/Teacher Rating Scale for Disruptive Behaviour Disorders (RS-DBD)[h] | | 8.5 | 7.2 | 43,238 |
| Child ADHD symptoms (inattention) age 8: Parent/Teacher Rating Scale for Disruptive Behaviour Disorders (RS-DBD)[i] | | 5.0 | 4.1 | 43,194 |
| Child ADHD symptoms (hyperactivity) age 8: Parent/Teacher Rating Scale for Disruptive Behaviour Disorders (RS-DBD)[j] | | 3.6 | 3.9 | 43,177 |
| **Categorical variables** | **Category** | | **%** | **N obs** |
| Child's sex | male | | 51.3 | 113,477 |
| | female | | 48.7 | |
| maternal educational qualifications | 9-year elementary education | | 2.8 | 101,020 |
| | Up to 2 years further education | | 5.2 | |
| | Further education: vocational | | 12.8 | |
| | Further education: general studies, sixth form | | 15.7 | |
| | Higher education: college/university, up to 4 years | | 40.9 | |
| | Higher education: college/university, over 4 years | | 22.6 | |
| paternal educational qualifications | 9-year elementary education | | 4.8 | 100,891 |
| | Up to 2 years further education | | 6.6 | |
| | Further education: vocational | | 26.2 | |
| | Further education: general studies, sixth form | | 13.2 | |
| | Higher education: college/university, up to 4 years | | 26.5 | |
| | Higher education: college/university, over 4 years | | 22.7 | |
| maternal parity at child's birth | 0 | | 44.0 | 113,691 |
| | 1 | | 35.9 | |
| | 2 | | 15.6 | |
| | 3 | | 3.4 | |
| | 4+ | | 1.1 | |

| | | | |
|---|---|---|---|
| Mother's marital status at birth | Married/registered partner | 95.5 | 113,691 |
| | single | 4.5 | |
| Mother's smoked during pregnancy | never | 49.9 | 101,373 |
| | Stopped before week 17 | 41.7 | |
| | Currently, sometimes | 2.8 | |
| | Currently, daily | 5.6 | |

[a]All participants in the birth registry file who had not withdrawn consent. [b]Possible range: 0-15. [c]Possible range: 0-24. [d]Possible range: 0-24. [e]Possible range: 0-24. [f]Possible range: 0-10. [g]Possible range: 0-54. [h]Possible range: 0-27. [i]Possible range: 0-27.

**Appendix 1 - Table 2: Descriptive statistics of analytic sample (N=40,949)[a], unimputed data**

| Continuous variables | | mean | SD | N obs |
|---|---|---|---|---|
| Maternal age at birth | | 30.2 | 4.4 | 40,949 |
| Paternal age at birth | | 32.6 | 5.1 | 40,945 |
| Maternal depressive/anxiety symptoms, based on 5 items from the Hopkins Symptoms Checklist-25 (SCL-25)[b] | | 1.2 | 1.9 | 39,647 |
| Paternal depressive/anxiety symptoms, based on 8 items from the Hopkins Symptoms Checklist-25 (SCL-25)[c] | | 1.1 | 2.1 | 38,050 |
| Maternal ADHD symptoms: adult ADHD self-report scale[d] | | 6.5 | 3.4 | 24,192 |
| Paternal ADHD symptoms: adult ADHD self-report scale[e] | | 8.2 | 3.1 | 16,348 |
| Maternal pre-pregnancy BMI (kg/m$^2$) | | 24.0 | 4.1 | 39,323 |
| Paternal BMI (kg/m$^2$) | | 25.9 | 3.2 | 39,405 |
| Child's BMI at age 8 (kg/m$^2$) | | 16.2 | 2.0 | 16,144 |
| Child depressive symptoms age 8: Short Mood and Feelings Questionnaire (SMFQ)[f] | | 1.8 | 2.4 | 18,747 |
| Child anxiety symptoms age 8: Screen for Child Anxiety Related Disorders (SCARED)[g] | | 1.0 | 1.2 | 18,834 |
| Child ADHD symptoms age 8: Parent/Teacher Rating Scale for Disruptive Behaviour Disorders (RS-DBD)[h] | | 8.4 | 7.1 | 18,813 |
| Child ADHD symptoms (inattention) age 8: Parent/Teacher Rating Scale for Disruptive Behaviour Disorders (RS-DBD)[i] | | 4.9 | 4.0 | 18,794 |
| Child ADHD symptoms (hyperactivity) age 8: Parent/Teacher Rating Scale for Disruptive Behaviour Disorders (RS-DBD)[j] | | 3.5 | 3.8 | 18,787 |
| **Categorical variables** | **Category** | | **%** | **N obs** |
| Child's sex | male | | 51.0 | 40,949 |
| | female | | 48.9 | |
| maternal educational qualifications | 9-year elementary education | | 2.0 | 39,569 |
| | Up to 2 years further education | | 4.1 | |
| | Further education: vocational | | 12.1 | |
| | Further education: general studies, sixth form | | 14.9 | |
| | Higher education: college/university, up to 4 years | | 42.8 | |
| | Higher education: college/university, over 4 years | | 24.8 | |
| paternal educational qualifications | 9-year elementary education | | 3.4 | 40,062 |
| | Up to 2 years further education | | 5.6 | |
| | Further education: vocational | | 25.1 | |
| | Further education: general studies, sixth form | | 12.9 | |
| | Higher education: college/university, up to 4 years | | 28.5 | |
| | Higher education: college/university, over 4 years | | 24.4 | |
| maternal parity at child's birth | 0 | | 46.8 | 40,949 |
| | 1 | | 35.7 | |
| | 2 | | 14.0 | |
| | 3 | | 2.7 | |
| | 4+ | | 0.7 | |

| | | | |
|---|---|---|---|
| Mother's marital status at birth | Married/registered partner | 97.4 | 40,949 |
| | single | 2.6 | |
| Mother's smoked during pregnancy | never | 51.1 | 40,118 |
| | Stopped before week 17 | 42.0 | |
| | Currently, sometimes | 2.4 | |
| | Currently, daily | 4.5 | |

[a]The reasons for exclusions and numbers in each case are shown in Appendix 1 - Figure 1. [b]Possible range: 0-15. [c]Possible range: 0-24. [d]Possible range: 0-24. [e]Possible range: 0-24. [f]Possible range: 0-10. [g]Possible range: 0-54. [h]Possible range: 0-27. [i]Possible range: 0-27.

**Appendix 1 - Table 3: BMI and symptoms of depression, anxiety, and ADHD at age 8 in MoBa, adult BMI PGS (N=40,949)[a]**

| Outcome | | Non-genetic estimate[b] Beta (per $5kg/m^2$) | CI | p | MR estimate[c] Beta (per $5kg/m^2$) | CI | p | Within-families MR estimate[c] Beta (per $5kg/m^2$) | CI | p |
|---|---|---|---|---|---|---|---|---|---|---|
| Depressive symptoms: standardized SMFQ[d] score | Child BMI[a] | 0.05 | 0.01,0.09 | 0.02 | 0.45 | 0.26,0.64 | <0.001 | 0.26 | -0.01,0.52 | 0.06 |
| | Mother's BMI | 0.05 | 0.03,0.07 | <0.001 | | | | 0.11 | 0.02,0.19 | 0.01 |
| | Father's BMI | 0.01 | -0.02,0.03 | 0.67 | | | | 0.02 | -0.09,0.13 | 0.71 |
| Anxiety symptoms: standardized SCARED[e] score | Child BMI | -0.07 | -0.11,-0.03 | 0.001 | -0.06 | -0.25,0.12 | 0.51 | 0.01 | -0.25,0.26 | 0.96 |
| | Mother's BMI | 0.01 | -0.01,0.03 | 0.47 | | | | -0.03 | -0.11,0.05 | 0.49 |
| | Father's BMI | -0.00 | -0.02,0.02 | 0.89 | | | | -0.02 | -0.13,0.09 | 0.72 |
| ADHD symptoms: standardized RS-DBD[f] score, ADHD items | Child BMI | -0.07 | -0.11,-0.03 | 0.001 | 0.35 | 0.17,0.53 | <0.001 | 0.36 | 0.09,0.63 | 0.009 |
| | Mother's BMI | 0.04 | 0.02,0.06 | <0.001 | | | | 0.00 | -0.08,0.09 | 0.97 |
| | Father's BMI | 0.02 | -0.00,0.04 | 0.10 | | | | -0.01 | -0.12,0.09 | 0.80 |
| ADHD-inattention symptoms: standardized RS-DBD[f] score, inattention items | Child BMI | -0.06 | -0.10,-0.02 | 0.006 | 0.32 | 0.14,0.49 | <0.001 | 0.38 | 0.12,0.65 | 0.005 |
| | Mother's BMI | 0.05 | 0.03,0.07 | <0.001 | | | | 0.01 | -0.08,0.09 | 0.86 |
| | Father's BMI | 0.02 | -0.00,0.05 | 0.06 | | | | -0.06 | -0.17,0.04 | 0.24 |
| ADHD-hyperactivity symptoms: standardized RS-DBD[f] score, hyperactivity items | Child BMI | -0.06 | -0.10,-0.02 | 0.002 | 0.31 | 0.13,0.49 | 0.001 | 0.27 | -0.00,0.54 | 0.05 |
| | Mother's BMI | 0.03 | 0.01,0.05 | 0.005 | | | | -0.00 | -0.09,0.08 | 0.92 |
| | Father's BMI | 0.01 | -0.01,0.04 | 0.32 | | | | 0.04 | -0.07,0.15 | 0.47 |

[a]Coefficients represent S.D. change in symptoms per $5kg/m^2$ increase in BMI. [b]Phenotypic models adjust for the child's sex and birth year, the mother's parity at the child's birth, and the mother's and father's: educational qualifications, depressive/anxiety and ADHD symptoms, and smoking status during pregnancy. They also adjust for the child's, mother's, and father's genotyping centre, genotyping chip, and first 20 principal components of ancestry. [c]Genetic models adjust for the child's sex and birth year and the child's, mother's, and father's genotyping centre, genotyping chip, and first 20 principal components of ancestry. [d]Short Mood and Feelings Questionnaire. [e]Screen for Child Anxiety Related Disorders. [f]Parent/Teacher Rating Scale for Disruptive Behaviour Disorders

**Appendix 1 - Table 4: BMI and symptoms of depression, anxiety, and ADHD at age 8 in MoBa, childhood body size PGS (N=40,949)[a]**

| | | Non-genetic estimate[b] | | | MR estimate[c] | | | Within-families MR estimate[c] | | |
|---|---|---|---|---|---|---|---|---|---|---|
| **Outcome** | | Beta (per 5kg/$m^2$) | CI | p | Beta (per 5kg/$m^2$) | CI | p | Beta (per 5kg/$m^2$) | CI | p |
| Depressive symptoms: standardized SMFQ[d] score | Child BMI[a] | 0.05 | 0.01,0.09 | 0.02 | 0.08 | -0.07,0.22 | 0.29 | 0.02 | -0.20,0.23 | 0.88 |
| | Mother's BMI | 0.05 | 0.03,0.07 | <0.001 | | | | 0.02 | -0.09,0.14 | 0.67 |
| | Father's BMI | 0.01 | -0.02,0.03 | 0.67 | | | | 0.05 | -0.10,0.19 | 0.51 |
| Anxiety symptoms: standardized SCARED[e] score | Child BMI | -0.07 | -0.11,-0.03 | 0.001 | -0.04 | -0.18,0.11 | 0.62 | 0.02 | -0.18,0.23 | 0.83 |
| | Mother's BMI | 0.01 | -0.01,0.03 | 0.47 | | | | -0.02 | -0.12,0.09 | 0.78 |
| | Father's BMI | -0.00 | -0.02,0.02 | 0.89 | | | | -0.06 | -0.19,0.08 | 0.42 |
| ADHD symptoms: standardized RS-DBD[f] score, ADHD items | Child BMI | -0.07 | -0.11,-0.03 | 0.001 | -0.07 | -0.21,0.07 | 0.35 | -0.03 | -0.22,0.17 | 0.80 |
| | Mother's BMI | 0.04 | 0.02,0.06 | <0.001 | | | | -0.03 | -0.12,0.07 | 0.62 |
| | Father's BMI | 0.02 | -0.00,0.04 | 0.10 | | | | -0.02 | -0.15,0.11 | 0.77 |
| ADHD-inattention symptoms: standardized RS-DBD[f] score, inattention items | Child BMI | -0.06 | -0.10,-0.02 | 0.006 | -0.04 | -0.18,0.11 | 0.63 | -0.05 | -0.25,0.14 | 0.59 |
| | Mother's BMI | 0.05 | 0.03,0.07 | <0.001 | | | | 0.03 | -0.07,0.13 | 0.61 |
| | Father's BMI | 0.02 | -0.00,0.05 | 0.06 | | | | -0.01 | -0.14,0.12 | 0.86 |
| ADHD-hyperactivity symptoms: standardized RS-DBD[f] score, hyperactivity items | Child BMI | -0.06 | -0.10,-0.02 | 0.002 | -0.08 | -0.22,0.05 | 0.24 | 0.01 | -0.20,0.22 | 0.95 |
| | Mother's BMI | 0.03 | 0.01,0.05 | 0.005 | | | | -0.07 | -0.18,0.03 | 0.18 |
| | Father's BMI | 0.01 | -0.01,0.04 | 0.32 | | | | -0.02 | -0.16,0.11 | 0.74 |

[a]Coefficients represent S.D. change in symptoms per 5kg/$m^2$ increase in BMI. [b]Phenotypic models adjust for the child's sex and birth year, the mother's parity at the child's birth, and the mother's and father's: educational qualifications, depressive/anxiety and ADHD symptoms, and smoking status during pregnancy. They also adjust for the child's, mother's, and father's genotyping centre, genotyping chip, and first 20 principal components of ancestry. [c]Genetic models adjust for the child's sex and birth year and the child's, mother's, and father's genotyping centre, genotyping chip, and first 20 principal components of ancestry. [d]Short Mood and Feelings Questionnaire. [e]Screen for Child Anxiety Related Disorders. [f]Parent/Teacher Rating Scale for Disruptive Behaviour Disorders

| **Appendix 1 - Table 5: Associations of phenotypes and polygenic scores within parental pairs*.** | | | | |
|---|---|---|---|---|
| *Phenotypes: regression of father's BMI, depressive symptoms, and ADHD symptoms on mother's phenotypes* | | | | |
| | *Father's:* BMI | *Father's:* Depressive symptoms | *Father's:* ADHD symptoms | |
| | Beta (95%CI), p | Beta (95%CI), p | Beta (95%CI), p | |
| *Mother's:* BMI | 0.23 (0.22,0.25) p<0.001 | 0.01 (-0.00,0.02) p=0.09 | 0.03 (0.02,0.05) p<0.001 | |
| *Mother's:* Depressive symptoms | 0.00 (-0.01,0.01), p=0.85 | 0.18 (0.16,0.20) p<0.001 | 0.10 (0.09,0.12), p<0.001 | |
| *Mother's:* ADHD symptoms | 0.01 (-0.02,0.01) p=0.32 | 0.05 (0.05,0.06) p<0.001 | 0.11 (0.09,0.13) p<0.001 | |
| *Polygenic scores: regression of father's PGS for BMI, depression, and ADHD: regression of father's PGS on mother's PGS* | | | | |
| | *Father's:* Adult BMI PGS | *Father's*: Childhood body size PGS | *Father's:* Depression PGS | *Father's:* ADHD PGS |
| | Beta (95%CI), p | Beta (95%CI), p | Beta (95%CI), p | Beta (95%CI) |
| *Mother's:* Adult BMI PGS | 0.01 (0.00,0.02), p=0.02 | 0.01 (0.00,0.02), p=0.008 | -0.00 (-0.01,0.01), p=0.62 | -0.00 (-0.01,0.01), p=0.49 |
| *Mother's:* Childhood body size PGS | 0.01 (-0.00,0.02), p=0.10 | 0.01 (-0.00,0.02), p=0.15 | 0.00 (-0.00,0.01), p=0.33 | 0.01 (0.00,0.02), p=0.03 |
| *Mother's:* Depression PGS | -0.01 (-0.02-0.00), p=0.11 | -0.00 (-0.01,0.01), p=0.76 | -0.00 (-0.01,0.01), p=0.44 | -0.00 (-0.01,0.01), p=0.47 |
| *Mother's:* ADHD PGS | -0.00 (-0.01,0.01), p=0.58 | 0.01 (-0.00,0.02), p=0.24 | -0.00 (-0.01,0.01), p=0.42 | -0.00 (-0.01,0.01), p=0.93 |
| *all models adjusted for the first 20 principal components of ancestry, genotyping centre and genotyping chip of the mother and father. | | | | |

**Appendix 1 - Table 6: BMI and symptoms of depression, anxiety, and ADHD at age 8 in MoBa, adult BMI PGS, log-transformed outcomes (N=40,949)[a]**

| | | **Non-genetic estimate[b]** | | | **MR estimate[c]** | | | **Within-families MR estimate[c]** | | |
|---|---|---|---|---|---|---|---|---|---|---|
| **Outcome** | | Beta (per 5kg/m²) | CI | p | Beta (per 5kg/m²) | CI | p | Beta (per 5kg/m²) | CI | p |
| Depressive symptoms: log-transformed SMFQ[d] score | Child BMI[a] | 0.03 | 0.01,0.06 | 0.02 | 0.31 | 0.18,0.45 | <0.001 | 0.16 | -0.04,0.35 | 0.12 |
| | Mother's BMI | 0.04 | 0.03,0.05 | <0.001 | | | | 0.08 | 0.02,0.14 | 0.01 |
| | Father's BMI | 0.01 | -0.01,0.02 | 0.56 | | | | 0.03 | -0.05,0.10 | 0.52 |
| Anxiety symptoms: log-transformed SCARED[e] score | Child BMI | -0.03 | -0.05,-0.01 | 0.001 | -0.04 | -0.14,0.05 | 0.38 | -0.00 | -0.14,0.13 | 0.97 |
| | Mother's BMI | 0.00 | -0.01,0.01 | 0.60 | | | | -0.02 | -0.06,0.02 | 0.35 |
| | Father's BMI | -0.00 | -0.01,0.01 | 1.00 | | | | -0.01 | -0.06,0.05 | 0.79 |
| ADHD symptoms: log-transformed RS-DBD[f] score | Child BMI | -0.05 | -0.08,-0.02 | 0.002 | 0.27 | 0.13,0.40 | <0.001 | 0.28 | 0.08,0.49 | 0.006 |
| | Mother's BMI | 0.03 | 0.02,0.05 | <0.001 | | | | 0.00 | -0.06,0.07 | 0.90 |
| | Father's BMI | 0.02 | -0.00,0.03 | 0.07 | | | | -0.02 | -0.10,0.06 | 0.66 |
| ADHD-inattention symptoms: log-transformed RS-DBD[f] score, inattention items | Child BMI | -0.04 | -0.07,-0.01 | 0.007 | 0.22 | 0.09,0.34 | 0.001 | 0.29 | 0.10,0.47 | 0.003 |
| | Mother's BMI | 0.03 | 0.02,0.05 | <0.001 | | | | 0.00 | -0.06,0.07 | 0.92 |
| | Father's BMI | 0.02 | 0.00,0.03 | 0.05 | | | | -0.06 | -0.14,0.02 | 0.13 |
| ADHD-hyperactivity symptoms: log-transformed RS-DBD[f] score, hyperactivity items | Child BMI | -0.05 | -0.08,-0.01 | 0.004 | 0.24 | 0.10,0.39 | 0.001 | 0.18 | -0.04,0.40 | 0.10 |
| | Mother's BMI | 0.02 | 0.00,0.04 | 0.01 | | | | 0.00 | -0.07,0.07 | 1.00 |
| | Father's BMI | 0.01 | -0.01,0.03 | 0.25 | | | | 0.05 | -0.04,0.14 | 0.29 |

[a]Coefficients represent change in symptoms, log-transformed after adding 1, per 5kg/m² increase in BMI. [b]Phenotypic models adjust for the child's sex and birth year, the mother's parity at the child's birth, and the mother's and father's: educational qualifications, depressive/anxiety and ADHD symptoms, and smoking status during pregnancy. They also adjust for the child's, mother's, and father's genotyping centre, genotyping chip, and first 20 principal components of ancestry. [c]Genetic models adjust for the child's sex and birth year and the child's, mother's, and father's genotyping centre, genotyping chip, and first 20 principal components of ancestry. [d]Short Mood and Feelings Questionnaire. [e]Screen for Child Anxiety Related Disorders. [f]Parent/Teacher Rating Scale for Disruptive Behaviour Disorders.

**Appendix 1 - Table 7: BMI and symptoms of depression, anxiety, and ADHD at age 8 in MoBa, childhood body size PGS, log-transformed outcomes (N=40,949)[a]**

| | | Non-genetic estimate[b] | | | MR estimate[c] | | | Within-families MR estimate[c] | | |
|---|---|---|---|---|---|---|---|---|---|---|
| Outcome | | Beta (per 5kg/m$^2$) | CI | p | Beta (per 5kg/m$^2$) | CI | p | Beta (per 5kg/m$^2$) | CI | p |
| Depressive symptoms: log-transformed SMFQ[d] score | Child BMI[a] | 0.03 | 0.01,0.06 | 0.02 | 0.07 | -0.04,0.17 | 0.20 | 0.02 | -0.14,0.17 | 0.84 |
| | Mother's BMI | 0.04 | 0.03,0.05 | <0.001 | | | | 0.02 | -0.06,0.10 | 0.67 |
| | Father's BMI | 0.01 | -0.01,0.02 | 0.56 | | | | 0.05 | -0.06,0.15 | 0.39 |
| Anxiety symptoms: log-transformed SCARED[e] score | Child BMI | -0.03 | -0.05,-0.01 | 0.001 | -0.03 | -0.10,0.05 | 0.49 | 0.01 | -0.10,0.12 | 0.89 |
| | Mother's BMI | 0.00 | -0.01,0.01 | 0.60 | | | | -0.01 | -0.07,0.04 | 0.69 |
| | Father's BMI | -0.00 | -0.01,0.01 | 1.00 | | | | -0.03 | -0.10,0.04 | 0.43 |
| ADHD symptoms: log-transformed RS-DBD[f] score | Child BMI | -0.05 | -0.08,-0.02 | 0.002 | -0.06 | -0.17,0.05 | 0.30 | -0.03 | -0.19,0.13 | 0.70 |
| | Mother's BMI | 0.03 | 0.02,0.05 | <0.001 | | | | -0.02 | -0.10,0.06 | 0.68 |
| | Father's BMI | 0.02 | -0.00,0.03 | 0.07 | | | | -0.01 | -0.11,0.09 | 0.79 |
| ADHD-inattention symptoms: log-transformed RS-DBD[f] score, inattention items | Child BMI | -0.04 | -0.07,-0.01 | 0.007 | -0.03 | -0.14,0.07 | 0.52 | -0.05 | -0.19,0.10 | 0.52 |
| | Mother's BMI | 0.03 | 0.02,0.05 | <0.001 | | | | 0.02 | -0.06,0.09 | 0.64 |
| | Father's BMI | 0.02 | 0.00,0.03 | 0.05 | | | | -0.01 | -0.10,0.09 | 0.87 |
| ADHD-hyperactivity symptoms: log-transformed RS-DBD[f] score, hyperactivity items | Child BMI | -0.05 | -0.08,-0.01 | 0.004 | -0.07 | -0.19,0.04 | 0.21 | -0.00 | -0.18,0.17 | 0.98 |
| | Mother's BMI | 0.02 | 0.00,0.04 | 0.01 | | | | -0.06 | -0.15,0.03 | 0.22 |
| | Father's BMI | 0.01 | -0.01,0.03 | 0.25 | | | | -0.02 | -0.13,0.09 | 0.73 |

[a]Coefficients represent change in symptoms, log-transformed after adding 1, per 5kg/m$^2$ increase in BMI. [b]Phenotypic models adjust for the child's sex and birth year, the mother's parity at the child's birth, and the mother's and father's: educational qualifications, depressive/anxiety and ADHD symptoms, and smoking status during pregnancy. They also adjust for the child's, mother's, and father's genotyping centre, genotyping chip, and first 20 principal components of ancestry. [c]Genetic models adjust for the child's sex and birth year and the child's, mother's, and father's genotyping centre, genotyping chip, and first 20 principal components of ancestry. [d]Short Mood and Feelings Questionnaire. [e]Screen for Child Anxiety Related Disorders. [f]Parent/Teacher Rating Scale for Disruptive Behaviour Disorders.

**Appendix 1 - Table 8: Robustness checks based on SNP-specific associations with child's BMI[a] and outcomes: SNPs in adult BMI polygenic score**

| | **Inverse-variance weighted** | | **MR-Egger: slope** | | **MR-Egger: intercept** | | **MR-Median** | | **MR-Modal** | |
|---|---|---|---|---|---|---|---|---|---|---|
| *For classic MR models* | Beta | p | Beta | p | Beta | p | Beta | p | Beta | p |
| Depressive symptoms: standardized SMFQ[b] score | 0.12 | <0.001 | 0.09 | 0.18 | 0.00 | 0.56 | 0.11 | <0.001 | 0.07 | 0.42 |
| Anxiety symptoms: standardized SCARED[c] score | -0.02 | 0.13 | -0.05 | 0.45 | 0.00 | 0.61 | -0.01 | 0.87 | 0.01 | 0.92 |
| ADHD symptoms: standardized RS-DBD[d] score | 0.10 | <0.001 | 0.02 | 0.77 | 0.00 | 0.19 | 0.09 | 0.01 | 0.07 | 0.40 |
| ADHD symptoms (inattention): standardized RS-DBD[d] score | 0.09 | <0.001 | -0.02 | 0.73 | 0.00 | 0.07 | 0.09 | 0.01 | -0.02 | 0.81 |
| ADHD symptoms (hyperactivity): standardized RS-DBD[d] score | 0.08 | <0.001 | 0.05 | 0.40 | 0.00 | 0.61 | 0.08 | 0.02 | 0.05 | 0.55 |
| | **Inverse-variance weighted** | | **MR-Egger: slope** | | **MR-Egger: intercept** | | **MR-Median** | | **MR-Modal** | |
| *For within-families MR models* | Beta | p | Beta | p | Beta | p | Beta | p | Beta | p |
| Depressive symptoms: standardized SMFQ[b] score | 0.06 | <0.001 | 0.04 | 0.67 | 0.00 | 0.77 | 0.07 | 0.16 | 0.02 | 0.83 |
| Anxiety symptoms: standardized SCARED[c] score | 0.00 | 1.00 | 0.05 | 0.58 | 0.00 | 0.57 | 0.03 | 0.57 | 0.03 | 0.76 |
| ADHD symptoms: standardized RS-DBD[d] score | 0.09 | <0.001 | 0.07 | 0.41 | 0.00 | 0.80 | 0.09 | 0.07 | 0.13 | 0.24 |
| ADHD symptoms (inattention): standardized RS-DBD[d] score | 0.10 | <0.001 | 0.00 | 0.97 | 0.00 | 0.21 | 0.10 | 0.03 | 0.05 | 0.63 |
| ADHD symptoms (hyperactivity): standardized RS-DBD[d] score | 0.07 | <0.001 | 0.13 | 0.13 | 0.00 | 0.44 | 0.07 | 0.14 | 0.06 | 0.56 |

[a]In the main analyses, all coefficients are expressed in terms of S.D. change in symptoms per 5kg/m$^2$ increase in BMI. In robustness checks, SNP-exposure associations were taken directly from the relevant GWAS. Coefficients above for BMI-outcome associations are therefore on the scale of kg/m$^2$. [b]Short Mood and Feelings Questionnaire. [c]Screen for Child Anxiety Related Disorders. [d]Parent/Teacher Rating Scale for Disruptive Behaviour Disorders.

**Appendix 1 - Table 9: Robustness checks based on SNP-specific associations with child's BMI[a] and outcomes: SNPs in childhood body size polygenic score**

| | **Inverse-variance weighted** | | **MR-Egger: slope** | | **MR-Egger: intercept** | | **MR-Median** | | **MR-Modal** | |
|---|---|---|---|---|---|---|---|---|---|---|
| *For classic MR models* | Beta | p | Beta | p | Beta | p | Beta | p | Beta | p |
| Depressive symptoms: standardized SMFQ[b] score | 0.06 | 0.03 | 0.10 | 0.30 | 0.00 | 0.57 | 0.05 | 0.52 | 0.04 | 0.67 |
| Anxiety symptoms: standardized SCARED[c] score | -0.02 | 0.40 | 0.01 | 0.90 | 0.00 | 0.66 | -0.02 | 0.74 | -0.01 | 0.95 |
| ADHD symptoms: standardized RS-DBD[d] score | -0.04 | 0.14 | -0.02 | 0.85 | 0.00 | 0.75 | -0.06 | 0.36 | -0.05 | 0.65 |
| ADHD symptoms (inattention): standardized RS-DBD[d] score | -0.02 | 0.47 | -0.04 | 0.64 | 0.00 | 0.81 | -0.08 | 0.28 | -0.08 | 0.42 |
| ADHD symptoms (hyperactivity): standardized RS-DBD[d] score | -0.05 | 0.05 | 0.01 | 0.90 | 0.00 | 0.42 | -0.05 | 0.48 | -0.04 | 0.68 |
| | **Inverse-variance weighted** | | **MR-Egger: slope** | | **MR-Egger: intercept** | | **MR-Median** | | **MR-Modal** | |
| *For within-families MR models* | Beta | p | Beta | p | Beta | p | Beta | p | Beta | p |
| Depressive symptoms: standardized SMFQ[b] score | 0.02 | 0.59 | 0.05 | 0.70 | 0.00 | 0.75 | 0.03 | 0.76 | 0.04 | 0.79 |
| Anxiety symptoms: standardized SCARED[c] score | 0.02 | 0.60 | 0.07 | 0.64 | 0.00 | 0.69 | 0.04 | 0.66 | 0.05 | 0.74 |
| ADHD symptoms: standardized RS-DBD[d] score | -0.02 | 0.53 | 0.09 | 0.49 | 0.00 | 0.35 | -0.01 | 0.94 | 0.03 | 0.85 |
| ADHD symptoms (inattention): standardized RS-DBD[d] score | -0.04 | 0.28 | 0.06 | 0.65 | 0.00 | 0.41 | -0.04 | 0.68 | 0.01 | 0.95 |
| ADHD symptoms (hyperactivity): standardized RS-DBD[d] score | 0.00 | 0.98 | 0.11 | 0.43 | 0.00 | 0.39 | 0.02 | 0.80 | 0.06 | 0.67 |

[a]In the main analyses, all coefficients are expressed in terms of S.D. change in symptoms per 5kg/m$^2$ increase in BMI. In robustness checks, SNP-exposure associations were taken directly from the relevant GWAS. Coefficients above for BMI-outcome associations are therefore on the scale of kg/m$^2$. [b]Short Mood and Feelings Questionnaire. [c]Screen for Child Anxiety Related Disorders. [d]Parent/Teacher Rating Scale for Disruptive Behaviour Disorders.

**Appendix 1 - Table 10: BMI and symptoms of depression, anxiety, and ADHD at age 8 in MoBa, adult BMI PGS, complete-case analysis[a]**

| | | **Non-genetic estimate[b]** | | | **MR estimate[c]** | | | **Within-families MR estimate[c]** | | |
|---|---|---|---|---|---|---|---|---|---|---|
| **Outcome** | | Beta (per 5kg/m$^2$) | CI | p | Beta (per 5kg/m$^2$) | CI | p | Beta (per 5kg/m$^2$) | CI | p |
| Depressive symptoms: standardized SMFQ[d] score. N=5158 | Child BMI[a] | 0.08 | 0.00,0.15 | 0.05 | 0.38 | 0.00,0.77 | 0.05 | 0.16 | -0.35,0.66 | 0.55 |
| | Mother's BMI | 0.07 | 0.03,0.11 | <0.001 | | | | 0.10 | -0.06,0.26 | 0.21 |
| | Father's BMI | 0.01 | -0.04,0.05 | 0.79 | | | | 0.04 | -0.15,0.23 | 0.66 |
| Anxiety symptoms: standardized SCARED[e] score. N=5177 | Child BMI | -0.04 | -0.12,0.03 | 0.26 | -0.15 | -0.54,0.24 | 0.45 | -0.07 | -0.62,0.48 | 0.80 |
| | Mother's BMI | 0.02 | -0.02,0.06 | 0.26 | | | | 0.06 | -0.11,0.22 | 0.48 |
| | Father's BMI | 0.01 | -0.04,0.05 | 0.69 | | | | -0.13 | -0.34,0.08 | 0.24 |
| ADHD symptoms: standardized RS-DBD[f] score, ADHD items. N=5174 | Child BMI | -0.03 | -0.11,0.04 | 0.38 | 0.41 | -0.00,0.83 | 0.05 | 0.54 | 0.01,1.08 | 0.04 |
| | Mother's BMI | 0.09 | 0.05,0.12 | <0.001 | | | | -0.01 | -0.16,0.15 | 0.93 |
| | Father's BMI | -0.01 | -0.05,0.03 | 0.60 | | | | -0.09 | -0.29,0.12 | 0.41 |
| ADHD-inattention symptoms: standardized RS-DBD[f] score, inattention items. N=5171 | Child BMI | -0.02 | -0.10,0.05 | 0.55 | 0.44 | 0.04,0.85 | 0.03 | 0.73 | 0.20,1.25 | 0.01 |
| | Mother's BMI | 0.09 | 0.05,0.12 | <0.001 | | | | -0.02 | -0.18,0.14 | 0.84 |
| | Father's BMI | -0.00 | -0.05,0.04 | 0.83 | | | | -0.18 | -0.39,0.03 | 0.09 |
| ADHD-hyperactivity symptoms: standardized RS-DBD[f] score, hyperactivity items. N=5167 | Child BMI | -0.04 | -0.11,0.03 | 0.29 | 0.28 | -0.13,0.70 | 0.18 | 0.21 | -0.32,0.75 | 0.43 |
| | Mother's BMI | 0.07 | 0.03,0.11 | 0.001 | | | | 0.01 | -0.15,0.17 | 0.87 |
| | Father's BMI | -0.01 | -0.06,0.03 | 0.51 | | | | 0.03 | -0.17,0.24 | 0.75 |

[a]Coefficients represent S.D. change in symptoms per 5kg/m$^2$ increase in BMI. [b]Phenotypic models adjust for the child's sex and birth year, the mother's parity at the child's birth, and the mother's and father's: educational qualifications, depressive/anxiety and ADHD symptoms, and smoking status during pregnancy. They also adjust for the child's, mother's, and father's genotyping centre, genotyping chip, and first 20 principal components of ancestry. [c]Genetic models adjust for the child's sex and birth year and the child's, mother's, and father's genotyping centre, genotyping chip, and first 20 principal components of ancestry. [e]Screen for Child Anxiety Related Disorders. [f]Parent/Teacher Rating Scale for Disruptive Behaviour Disorders.

**Appendix 1 - Table 11: BMI and symptoms of depression, anxiety, and ADHD at age 8 in MoBa, childhood body size PGS, complete-case analysis[a]**

| Outcome | | Non-genetic estimate[b] Beta (per 5kg/m²) | CI | p | MR estimate[c] Beta (per 5kg/m²) | CI | p | Within-families MR estimate[c] Beta (per 5kg/m²) | CI | p |
|---|---|---|---|---|---|---|---|---|---|---|
| Depressive symptoms: | Child BMI[a] | 0.08 | 0.00,0.15 | 0.05 | 0.13 | -0.15,0.42 | 0.37 | 0.10 | -0.34,0.54 | 0.65 |
| standardized SMFQ[d] score | Mother's BMI | 0.07 | 0.03,0.11 | <0.001 | | | | -0.06 | -0.26,0.14 | 0.57 |
| N=5158 | Father's BMI | 0.01 | -0.04,0.05 | 0.79 | | | | 0.10 | -0.20,0.40 | 0.52 |
| Anxiety symptoms: | Child BMI | -0.04 | -0.12,0.03 | 0.26 | -0.03 | -0.32,0.27 | 0.85 | 0.06 | -0.37,0.48 | 0.79 |
| standardized SCARED[e] score | Mother's BMI | 0.02 | -0.02,0.06 | 0.26 | | | | 0.02 | -0.19,0.24 | 0.85 |
| N=5177 | Father's BMI | 0.01 | -0.04,0.05 | 0.69 | | | | -0.13 | -0.43,0.18 | 0.42 |
| ADHD symptoms: | Child BMI | -0.03 | -0.11,0.04 | 0.38 | 0.00 | -0.29,0.30 | 0.98 | 0.13 | -0.31,0.56 | 0.57 |
| standardized RS-DBD[f] score, | Mother's BMI | 0.09 | 0.05,0.12 | <0.001 | | | | -0.05 | -0.25,0.15 | 0.61 |
| ADHD items | Father's BMI | -0.01 | -0.05,0.03 | 0.60 | | | | -0.09 | -0.40,0.22 | 0.56 |
| N=5174 | | | | | | | | | | |
| ADHD-inattention symptoms: | Child BMI | -0.02 | -0.10,0.05 | 0.55 | -0.00 | -0.29,0.29 | 0.99 | 0.13 | -0.29,0.55 | 0.55 |
| standardized RS-DBD[f] score, | Mother's BMI | 0.09 | 0.05,0.12 | <0.001 | | | | 0.04 | -0.17,0.24 | 0.72 |
| inattention items | Father's BMI | -0.00 | -0.05,0.04 | 0.83 | | | | -0.20 | -0.51,0.11 | 0.20 |
| N=5171 | | | | | | | | | | |
| ADHD-hyperactivity | Child BMI | -0.04 | -0.11,0.03 | 0.29 | 0.00 | -0.30,0.30 | 0.99 | 0.10 | -0.35,0.55 | 0.67 |
| symptoms: standardized RS- | Mother's BMI | 0.07 | 0.03,0.11 | 0.001 | | | | -0.14 | -0.34,0.06 | 0.18 |
| DBD[f] score, hyperactivity | Father's BMI | -0.01 | -0.06,0.03 | 0.51 | | | | 0.03 | -0.28,0.34 | 0.84 |
| items | | | | | | | | | | |
| N=5167 | | | | | | | | | | |

[a]Coefficients represent S.D. change in symptoms per 5kg/m² increase in BMI. [b]Phenotypic models adjust for the child's sex and birth year, the mother's parity at the child's birth, and the mother's and father's: educational qualifications, depressive/anxiety and ADHD symptoms, and smoking status during pregnancy. They also adjust for the child's, mother's, and father's genotyping centre, genotyping chip, and first 20 principal components of ancestry. [c]Genetic models adjust for the child's sex and birth year and the child's, mother's, and father's genotyping centre, genotyping chip, and first 20 principal components of ancestry. [d]Short Mood and Feelings Questionnaire. [e]Screen for Child Anxiety Related Disorders. [f]Parent/Teacher Rating Scale for Disruptive Behaviour Disorders.

**Appendix 1 - Table 12: Multivariable-adjusted associations[a] of BMI quintiles with symptoms of depression, anxiety, and ADHD at age 8 in MoBa**

| | Depressive symptoms: standardized SMFQ[b] score | | Anxiety symptoms: standardized SCARED[c] score | | ADHD symptoms: standardized RS-DBD[d] score, ADHD items | | ADHD-inattention symptoms: standardized RS-DBD[d] score, inattention items | | ADHD-hyperactivity symptoms: standardized RS-DBD[d] score, hyperactivity items | |
|---|---|---|---|---|---|---|---|---|---|---|
| **BMI quintile** | Beta[e] (95%CI) | p | Beta[e] (95%CI) | p | Beta[e] (95%CI) | p | Beta[e] (95%CI) | p | Beta[e] (95%CI) | p |
| 1 | 0.00 (-0.04,0.04) | 0.99 | 0.04 (0.00,0.09) | 0.04 | 0.05 (0.01,0.09) | 0.01 | 0.05 (0.01,0.09) | 0.02 | 0.04 (-0.00,0.08) | 0.05 |
| 2 | -0.01 (-0.05,0.03) | 0.70 | 0.02 (-0.03,0.06) | 0.47 | 0.02 (-0.02,0.06) | 0.29 | 0.02 (-0.02,0.06) | 0.38 | 0.02 (-0.02,0.06) | 0.30 |
| 3 (ref) | 1 | | 1 | | 1 | | 1 | | 1 | |
| 4 | 0.01 (-0.03,0.05) | 0.62 | -0.03 (-0.07,0.01) | 0.16 | -0.01 (-0.04,0.03) | 0.66 | -0.01 (-0.05,0.02) | 0.54 | -0.00 (-0.04,0.03) | 0.87 |
| 5 | 0.04 (0.00,0.08) | 0.03 | -0.03 (-0.07,0.01) | 0.09 | -0.02 (-0.06,0.02) | 0.27 | -0.01 (-0.05,0.03) | 0.51 | -0.03 (-0.06,0.01) | 0.20 |

[a]Models adjust for the child's sex and birth year, the mother's parity at the child's birth, and the mother's and father's: educational qualifications, depressive/anxiety and ADHD symptoms, and smoking status during pregnancy. They also adjust for the child's, mother's, and father's genotyping centre, genotyping chip, and first 20 principal components of ancestry. [b]Short Mood and Feelings Questionnaire. [c]Screen for Child Anxiety Related Disorders. [d]Parent/Teacher Rating Scale for Disruptive Behaviour Disorders. [e]Coefficients represent S.D. difference in symptoms between quintiles of child's BMI.

**Appendix 1 - Table 13: BMI and symptoms of depression, anxiety, and ADHD at age 8 in MoBa, adult BMI PGS, genetic models adjusted for parental education (N=40,949)[a]**

| Outcome | | MR estimate[b] Beta (per 5kg/$m^2$) | CI | p | Within-families MR estimate[b] Beta (per 5kg/$m^2$) | CI | p |
|---|---|---|---|---|---|---|---|
| Depressive symptoms: standardized SMFQ[c] score | Child BMI[a] | 0.38 | 0.19,0.58 | <0.001 | 0.26 | -0.01,0.53 | 0.06 |
| | Mother's BMI | | | | 0.09 | -0.00,0.17 | 0.05 |
| | Father's BMI | | | | -0.00 | -0.11,0.11 | 0.97 |
| Anxiety symptoms: standardized SCARED[d] score | Child BMI | -0.08 | -0.26,0.11 | 0.43 | 0.01 | -0.25,0.27 | 0.95 |
| | Mother's BMI | | | | -0.04 | -0.12,0.05 | 0.40 |
| | Father's BMI | | | | -0.03 | -0.14,0.08 | 0.64 |
| ADHD symptoms: standardized RS-DBD[e] score, ADHD items | Child BMI | 0.27 | 0.09,0.45 | 0.003 | 0.37 | 0.10,0.64 | 0.007 |
| | Mother's BMI | | | | -0.03 | -0.12,0.06 | 0.52 |
| | Father's BMI | | | | -0.05 | -0.16,0.06 | 0.37 |
| ADHD-inattention symptoms: standardized RS-DBD[e] score, inattention items | Child BMI | 0.25 | 0.07,0.42 | 0.007 | 0.39 | 0.13,0.66 | 0.004 |
| | Mother's BMI | | | | -0.02 | -0.11,0.07 | 0.64 |
| | Father's BMI | | | | -0.10 | -0.21,0.01 | 0.08 |
| ADHD-hyperactivity symptoms: standardized RS-DBD[e] score, hyperactivity items | Child BMI | 0.24 | 0.06,0.42 | 0.009 | 0.27 | 0.00,0.55 | 0.05 |
| | Mother's BMI | | | | -0.03 | -0.12,0.06 | 0.49 |
| | Father's BMI | | | | 0.01 | -0.10,0.12 | 0.85 |

[a]Coefficients represent S.D. change in symptoms per 5kg/$m^2$ increase in BMI. [b]Models adjust for the child's sex and birth year and the child's, mother's, and father's genotyping centre, genotyping chip, and first 20 principal components of ancestry. [c]Short Mood and Feelings Questionnaire. [d]Screen for Child Anxiety Related Disorders. [e]Parent/Teacher Rating Scale for Disruptive Behaviour Disorders.

**Appendix 1 - Table 14: BMI and symptoms of depression, anxiety, and ADHD at age 8 in MoBa, childhood body size PGS, genetic models adjusted for parental education (N=40,949)[a]**

| Outcome | | MR estimate[b] Beta (per 5kg/m$^2$) | CI | p | Within-families MR estimate[b] Beta (per 5kg/m$^2$) | CI | p |
|---|---|---|---|---|---|---|---|
| Depressive symptoms: standardized SMFQ[c] score | Child BMI[a] | 0.07 | -0.07,0.22 | 0.33 | 0.01 | -0.20,0.23 | 0.90 |
| | Mother's BMI | | | | 0.02 | -0.10,0.13 | 0.76 |
| | Father's BMI | | | | 0.05 | -0.09,0.20 | 0.48 |
| Anxiety symptoms: standardized SCARED[d] score | Child BMI | -0.03 | -0.18,0.11 | 0.65 | 0.03 | -0.18,0.23 | 0.81 |
| | Mother's BMI | | | | -0.02 | -0.13,0.09 | 0.76 |
| | Father's BMI | | | | -0.06 | -0.19,0.08 | 0.44 |
| ADHD symptoms: standardized RS-DBD[e] score, ADHD items | Child BMI | -0.08 | -0.22,0.06 | 0.27 | -0.03 | -0.23,0.16 | 0.75 |
| | Mother's BMI | | | | -0.03 | -0.13,0.07 | 0.50 |
| | Father's BMI | | | | -0.02 | -0.15,0.11 | 0.79 |
| ADHD-inattention symptoms: standardized RS-DBD[e] score, inattention items | Child BMI | -0.05 | -0.20,0.10 | 0.52 | -0.06 | -0.25,0.14 | 0.57 |
| | Mother's BMI | | | | 0.02 | -0.08,0.12 | 0.74 |
| | Father's BMI | | | | -0.01 | -0.14,0.12 | 0.87 |
| ADHD-hyperactivity symptoms: standardized RS-DBD[e] score, hyperactivity items | Child BMI | -0.10 | -0.23,0.04 | 0.18 | -0.00 | -0.21,0.21 | 1.00 |
| | Mother's BMI | | | | -0.08 | -0.19,0.03 | 0.15 |
| | Father's BMI | | | | -0.02 | -0.15,0.12 | 0.77 |

[a]Coefficients represent S.D. change in symptoms per 5kg/m$^2$ increase in BMI. [b]Models adjust for the child's sex and birth year and the child's, mother's, and father's genotyping centre, genotyping chip, and first 20 principal components of ancestry. [c]Short Mood and Feelings Questionnaire. [d]Screen for Child Anxiety Related Disorders. [e]Parent/Teacher Rating Scale for Disruptive Behaviour Disorders.
